# The SARS-CoV-2 receptor-binding domain expressed in Pichia pastoris as a candidate vaccine antigen

**DOI:** 10.1101/2021.06.29.21259605

**Authors:** Miladys Limonta-Fernández, Glay Chinea-Santiago, Alejandro Miguel Martín-Dunn, Diamile Gonzalez-Roche, Monica Bequet-Romero, Gabriel Marquez-Perera, Isabel González-Moya, Camila Canaan-Haden-Ayala, Ania Cabrales-Rico, Luis Ariel Espinosa-Rodríguez, Yassel Ramos-Gómez, Ivan Andujar-Martínez, Luis Javier González-López, Mariela Perez de la Iglesia, Jesus Zamora-Sanchez, Otto Cruz-Sui, Gilda Lemos-Pérez, Gleysin Cabrera-Herrera, Jorge Valdes-Hernández, Eduardo Martinez-Diaz, Eulogio Pimentel-Vazquez, Marta Ayala-Avila, Gerardo Guillén-Nieto

## Abstract

1.

The effort to develop vaccines based on economically accessible technological platforms available by developing countries vaccine manufacturers is essential to extend the immunization to the whole world population and to achieve the desired herd immunity, necessary to end the COVID-19 pandemic. Here we report on the development of a SARS-CoV-2 receptor-binding domain (RBD) protein, expressed in yeast *Pichia pastoris.* The RBD was modified with addition of flexible N- and C-terminal amino acid extensions aimed to modulate the protein/protein interactions and facilitate protein purification. Fermentation with yeast extract culture medium yielded 30–40 mg/L. After purification by immobilized metal ion affinity chromatography and hydrophobic interaction chromatography, the RBD protein was characterized by mass-spectrometry, circular dichroism, and binding affinity to angiotensin-converting enzyme 2 (ACE2) receptor. The recombinant protein shows high antigenicity with convalescent human sera and also with sera from individuals vaccinated with the Pfizer-BioNTech mRNA or Sputnik V adenoviral-based vaccines. The RBD protein stimulates IFNγ, IL-2, IL-6, IL-4, and TNFα in mice secreting splenocytes from PBMC and lung CD3+ enriched cells. Immunogenicity studies with 50 µg of the recombinant RBD formulated with alum, induce high levels of binding antibodies in mice and non-human primates, assessed by ELISA plates covered with RBD protein expressed in HEK293T cells. The mouse sera inhibited the RBD binding to ACE2 receptor in an *in-vitro* test and show neutralization of SARS-CoV-2 infection of Vero E6 cells. These data suggest that the RBD recombinant protein expressed in yeast *P. pastoris* is suitable as a vaccine candidate against COVID-19.

**Highlights:**

- The RBD protein (C-RBD-H6 PP) is expressed with high purity in *P. pastoris*.
- Physico-chemical characterization confirms the right folding of the protein.
- The recombinant protein shows high antigenicity with sera from convalescents.
- The sera from animals inhibit the RBD-ACE2 binding and neutralize the virus.
- The C-RBD-H6 protein stimulates IFNγ, IL-2, IL-6, IL-4, and TNFα in mice.

## 1. Introduction

The pandemic caused by SARS-CoV-2 has demonstrated the need to modify the way that public health, the governmental role, and international cooperation in science and health have been functioning up to now. It will not be possible to end the pandemic if the desired herd immunity is not achieved and for that it is necessary to vaccinate the vast majority of the world’s population in the shortest possible time. It is not enough for industrialized countries to be vaccinated, so the contribution of the developing countries’ vaccine manufacturers is more than welcome.

Cuba is an example of a global vision of public health and stands out for the strength of its biotechnology industry where one of the main achievements is the production of the Hepatitis B recombinant vaccine in yeast *P. pastoris* since 1991 [1–3] and the inclusion in the national immunization program since 1992. As a result, the incidence of acute hepatitis B in the country has decreased down to less than 50 cases and acute hepatitis B in children under 5 years old has not been reported since the year 2000 [4].

Therefore, it is feasible to select this technological platform for the development of a vaccine candidate against SARS-CoV-2.

The mammalian cell expression systems as Chinese hamster ovary cells (CHO), baby hamster kidney cells (BHK21), human embryonic kidney cells (HEK293T), and mouse-derived myeloma cell lines (NS0, SP2/0) are preferred for the expression of complex proteins for its capacity of proper post-translational modifications equivalent to that of humans, even though the selection of this recombinant protein production platform is limited by its low growth and productivity, and high production cost due to the relevant nutrient requirement [5]. The potential viral contamination of culture medium has also limited the use of the mammalian cell expression systems in large-scale production [6].

*Pichia pastoris* is one of the biotechnological platforms for the production of recombinant proteins that overcome several limitations of bacteria as expression system, such as protein aggregation and misfolding, production of toxic lipopolysaccharides, lack of posttranslational modifications, and frequent protein degradation due to the presence of proteases [7]. N-glycans in yeast secreted proteins are highly mannosylated while mammalian proteins have hybrid sugar composition. *P. pastoris* uses methanol, an inductor of the strong and tightly regulated AOX1 promoter as an exclusive carbon source. Therefore methanol can be used to drive protein expression altogether that it is reached an efficient secretion of the recombinant proteins [8]. On the other hand, the advantages of protein production by *P. pastoris* lie in the fact that the mechanism of protein expression in these microorganisms is close to the ones in mammalian cells and include the right protein folding in the endoplasmic reticulum, protein secretion by Kex2 as signal peptidase and low contamination with host proteins due to its limited production of endogenous secretory proteins [9], which simplifies and makes the purification process more feasible. Other significant advantages of yeast include growth speed, and easy genetic manipulation allowing linearized foreign DNA to be inserted at high efficiency in a chromosome via cross recombination to generate stable cell lines [10].

The receptor-binding domain of SARS-CoV-2 is a glycosylated 25 kDa protein domain spanning residues N_331_-K_529_ of the spike protein, including eight cysteine residues forming four disulfide bonds. The domain contains two glycosylation sites (N_331_ and N_343_) and a central twisted antiparallel beta-sheet formed by five strands with secondary structure short helices and loops [11]. Although glycosylation plays a fundamental role in the immunogenicity and stability of the RBD protein, the variability of the composition and heterogeneity in size of the sugar chains may not influence the receptor binding capacity [12]. Probably, for this reason the glycosylation introduced by *P. pastoris* more distant from mammalian cells and therefore more distant to the humans could contribute to the protein immunogenicity. RBD mediates cell entry through ACE2 host receptor and the levels of RBD binding antibodies strongly correlate with neutralizing antibodies in patients. Even more, the neutralizing antibody kinetics in patients mirrored the kinetics of RBD antibody development [13]. Since RBD upon infection is the main target of neutralizing antibodies, it has become the focus of vaccine design.

The SARS-CoV-2 RBD has been expressed at high levels in *P. pastoris* as a suitable vaccine candidate against COVID-19 [8;14;15]. The comparison between RBD obtained from *P. pastoris* and HEK293T mammalian cells by CD and tryptophan fluorescence shows that proteins were properly folded as well as that they have similar temperature stability despite differences in glycosylation of both expression platforms.

Here, we report the design of an RBD protein vaccine candidate, its expression in *P. pastoris* yeast, protein purification, the physico-chemical characterization and the capacity of the protein to elicit ACE2 binding inhibition antibodies, and neutralizing response in mice and monkeys. Our approach differs from the previously reported expression of RBD in *P. pastoris* [8] in the inclusion of the RBD glycosylation sites maintaining the advantage of the glycosylation capacity of *P. pastoris*.

## 2. Materials and Methods

### 2.1. Biological reagents, protein codification and serum panels

Human and murine RBD and ACE2 receptor chimeric proteins (hFc-RBD, mFc-RBD, hFc-ACE2, mFc-ACE2) were supplied by the Center of Molecular Immunology (CIM, Havana, Cuba). All the chimeric proteins were purified by protein-A based purification of the supernatant of stable transduce HEK293T cells and eluted in PBS. hFc-RBD was conjugated with peroxidase (HRP) and henceforth named hFcRBD-HRP. H6-RBD was produced as *E. coli* inclusion bodies [16], while RBD-H6 and C-RBD-H6 HEK were secreted to the supernatant of stably transduced HEK293T. All three proteins were purified by IMAC and the final buffer was exchanged to PBS. N-terminal end segment (C), and C-terminal end six histidine tag (H6), added to the central RBD protein sequence in a different protein constructions are codified as C-RBD-H6. Acronyms PP and HEK next to the protein code refers to the expression system when *P. pastoris* or HEK293T cells where used.

Different human sera used as controls include sera from volunteers vaccinated with Pfizer/BioNtech or Gamaleya’s Sputnik V (Gam-COVID-Vac) vaccine, and sera from convalescent patients. All individuals gave their writing informed consent for the use of their serum.

### 2.2. Construction of Pichia pastoris strains expressing RBD from SARS-CoV-2 (C-RBD-H6 PP)

A sequence coding for residues 331-530 of the Spike protein of SARS-CoV-2 strain Wuhan-Hu-1 (NCBI Acc. No. YP_009724390) with the appropriate N- and C-terminal extensions was codon-optimized for *S. cerevisiae* using J-Cat [17] and cloned in-frame with the KEX2 cleavage site of the pre-pro MATα sequence of pPICZαA (Invitrogen, USA), placing it under transcriptional control of the *P. pastoris* AOX1 promoter. After sequence verification, a representative clone was used to transform *P. pastoris* strain X-33 [18], selecting transformants by plating in YPD agar at Zeocin™ concentrations of 100, 200, 400, and 800 µg/mL. Three transformants from the plate with the highest concentration of Zeocin™ were randomly picked, purified by further streaking on YPD-Zeocin™, and used to prepare small seed banks. The seed banks were used in turn to inoculate small-scale (50 mL) BMGY cultures [18] that were induced after 24 h of growth at 28 °C by the addition of 0.5 % methanol. Culture supernatants were analyzed at 96 h post-induction by SDS-PAGE and Western blotting with an anti-His6 antibody (Promega, USA).

### 2.3. Fermentation

Fermentation was carried out in a 75-liters Chemap fermenter (Germany) with a working volume of 50 L of fermentation medium containing per liter of culture 8.8 mL of 85 % phosphoric acid, 6.92 g MgSO_4_.7H_2_O, 1.23 g (NH_4_)_2_SO_4_, 16.77 g K_2_HPO_4_, 0.46 g CaCl_2_.2H_2_O, 32.3 mL of 98% glycerol, 4.61 g yeast extract, 4 mL histidine solution, 5 mL of 400X vitamin base solution (1 g L^−1^ myo-inositol, 0.8 g L^−1^ D-calcium pantothenate, 0.8 g L^−1^ thiamine hydrochloride, 0.8 g L^−1^ pyridoxol hydrochloride, 0.2 g L^−1^ nicotinic acid, 0.8 mg L^−1^ D (+) Biotin), and 1 mL of sterile-filtered trace 1000X element solution (6 g L^−1^ CuSO_4_.5H_2_O, 0.415 g L^−1^ KI, 3 g L^−1^ MnSO_4_.H_2_O, 1 g L^−1^ Na_2_MoO_4_.2H_2_O, 0.1 g L^−1^ H_3_BO_3_, 20 g L^−1^ ZnSO_4_.7H_2_O, 65 g L^−1^ FeSO_4_.7H_2_O, 10 mL. conc.H_2_SO_4_) was added after autoclaving [19]. After inoculation of the bioreactor with 4 L of pre-culture, the temperature was maintained at 30 °C and the pH at 4.75 by pumping in a liquid ammonia solution. When a dissolved oxygen peak or a widening of the pH peaks is observed, the fed-batch phase is started with glycerol 50 % at 540 mL/h for 2-3 h. After one hour, the temperature is lowered to 25 ° C and raises the pH to 5.5. When the 50 % glycerol increase is exhausted, 800 mL of methanol is added at maximum flow of the peristaltic pump. The induction phase with methanol is started at an initial flow of 240 mL/h, 4 h later the flow is increased to 380 mL/h, and then at 480 mL/h. This last flow is maintained until the end of fermentation (38-44 h).

### 2.4. Protein purification

After 48 h of fermentation in a yeast extract culture medium, the culture was harvested, and cells were removed by centrifugation retention time between 5 −10 min at 15000 rpm at 4 °C. The fermentation supernatant was filtered in tandem conditions from 8 μm – 3 μm – 0.45 μm using cellulose filters. The supernatant was concentrated with a tangential flow filtration system with 30 kDa Hydrosart® membrane (Sartorius, Germany), besides buffer exchanged was carried out against PBS buffer containing 5 mM of imidazole. The IMAC column (Chelating Sepharose™ FF, Cytiva) was equilibrated in the same buffer and the sample was loaded and sequentially washed by using 30 column volumes of PBS containing 10 mM and 20 mM imidazole. Elution was carried out by increasing the concentration of imidazole up to 250 mM in the equilibrium buffer. The eluted protein was applied to a RP C4 column (Tosohaas, Japan) with dimensions 50 mm in diameter by 250 mm long for a resin volume of 500 mL and with a particle size of 15 to 20 µm. The column is coupled to a Shimadzu model LC-20AP semi-preparative HPLC purification system. The column was equilibrated with 0.5 % TFA solution, and protein was eluted by using a linear gradient of 1 % TFA solution (RP solution A) and acetonitrile with 0.5 % TFA (RP solution B) from 32 to 45 % solution B in 40 minutes. The protein elutes between 35 and 37 % of solution B, approximately in one column volume. The fraction collected with the purified protein was concentrated by tangential flow filtration system with a 10 kDa Hydrosart® membrane and pooled aseptically using a 0.22 μm and stored at −20 °C.

### 2.5. ESI-MS analysis of the deglycosylated C-RBD-H6 PP protein

The volume equivalent to 100 μg of the protein dissolved in PBS (pH 7.2) was deglycosylated with 1μL of PNGase-F (500 units, New England Biolabs) in presence of 0.5 M guanidine hydrochloride and 5 mM N-ethylmaleimide during 2 h at 37 °C. An aliquot of 10 μg of deglycosylated protein was desalted by using ZipTip C18 (Millipore) and the elution was loaded into the metal-coated nanocapillary for ESI-MS analysis. The remaining deglycosylated protein was digested by using an in-solution buffer-free trypsin digestion protocol previously reported [20] and adapted to the analysis of SARS-CoV-2 RBD proteins by introducing some modifications that provide full-sequence coverage and detection of post-translational modifications in a single ESI-MS spectrum [16]. Other experimental conditions for ESI-MS analysis are similar to those reported previously [16].

### 2.6. Surface plasmon resonance experimental procedure

The interaction between mFc-ACE2 fusion protein and the recombinant C-RBD-H6 PP was monitored by SPR using a BIACORE X (GE Health-care) at 25 °C in a multi-cycle mode. Briefly, mFc-ACE2 was immobilized on a Protein A biosensor chip (GE Health-care) according to the manufacturer’s protocol through the flow cell 1 (FC1). The FC2 was used as the reference cell. The real-time response of the C-RBD-H6 PP over the immobilized mFc-ACE2 was recorded by duplicate in a concentration range from 15 to 2000 nM, at 10 µL/min flow rate for 120 s, while the dissociation took place for another 120 s. The running buffer was PBS (pH = 7.2). After each cycle the chip was regenerated using pH = 2.0 glycine buffer. The equilibrium dissociation constant (binding affinity, K_D_) was estimated with the BIAevaluation® software (GE Healthcare) using the Langmuir 1:1 interaction model. At least five curves were taken into account for kinetics calculations.

### 2.7. Structural Analysis by Circular Dichroism (CD) Spectroscopy

CD spectra were acquired in a Jasco J-1500 CD spectrometer (Jasco, Japan). All measurements were carried out at 24 °C, the far UV CD spectra were studied at 100 µg/ml protein concentration (a 10 fold dilution in water of the stock solution) using a 1mm quartz cuvette. The near UV CD spectra were studied at the protein concentration of the stock solution in 20 mM pH 7.4 Tris buffer using a quartz cuvette of 10 mm path length. The spectra of the corresponding solution were subtracted. The far UV CD spectra were further analyzed by the BeStSel method [21;22] to estimate the secondary structure content of the protein and the results were compared with the values derived from the 3D coordinates of the crystallographic structure of the spike protein of SARS-CoV-2 (PDB file 6yla) using DSSP method [23] implemented in Whatif program package [24].

### 2.8. Animals and immunization schedules

Three different animal species were used for evaluation of immunogenicity of the C-RBD-H6 PP protein: BALB/c mice, Sprague–Dawley (SD) rats, and African green monkeys (*Chlorocebus aethiops sabaeus*). Six- to eight-week-old female BALB/c mice, and male and female SD rats were used for the study and housed in the animal facility. The experimental protocols were approved by the Ethical Committee on Animal Experimentation of the Center for Genetic Engineering and Biotechnology (CIGB, Havana, Cuba) and the Center for Production of Laboratory Animals (CENPALAB, Bejucal, Cuba).

The immunogen content per 500 µL: 50 µg of C-RBD-H6 PP protein adjuvated with 0.3 mg of aluminum hydroxide gel (Alhydrogel ®) in phosphate buffer (0.28 mg of disodium hydrogen phosphate, 0.31 mg of sodium dihydrogen phosphate dihydrate, 4.25 mg of sodium chloride).

*BALB/c mice*: Immunogenicity in mice was evaluated using a three-dose schedule with a 50 µg dose by intraperitoneal route with a 7 and 14 days interval before the second and third doses respectively. Blood was collected a week after the first boost and 7 and 14 days after the second boost. The number of animals per group, age, and gender of the animals are described in individual experiments.

#### Sprague Dawley rats

Immunogenicity of the C-RBD-H6 PP protein was tested in 20 SD rats (10 male and 10 female) during chronic toxicology study, with a 9 µg dose administered by the intramuscular route once a week, for 10 consecutive weeks for a total of 90 µg. Animals were bled three days after the last dose.

#### *Chlorocebus aethiops sabaeus* non-human primates

NHP ages between 3 to 6 years and with 2-7 kg of weight were kept at the animal’s facility at the CENPALAB. A total of 20 NHP were randomly assigned to 3 groups including the placebo (2 animals/gender, total 4), the low dose (50 µg, 3 animals/gender, total 6), and the high dose (100 µg, 5 animals/gender, total 10). Seven days post-1st and 2nd boosting and after overnight fasting, monkeys were sedated by intramuscular injection of ketamine hydrochloride (10 mg/kg) and bled from the femoral vein.

Specific anti-RBD titers and the inhibition of its interaction with ACE2 receptor were evaluated using ELISA. Live SARS-CoV-2 neutralization was assessed using a microneutralization assay.

### 2.9. Serum antibodies evaluation

#### Antibody detection by ELISA

The reactivity of sera from immunized animals was determined by ELISA. Briefly, 0.25 µg of RBD protein produced in HEK293T cells (Center for Molecular Immunology, Havana) was used to coat 96-well microtiter plates (Corning Costar, Acton, MA) in 0.1 M sodium carbonate buffer (pH 9.6) at 4 °C overnight. After the plates were blocked with 2 % skim milk, 0.05 % Tween 20, serially diluted mouse, rat or NHP sera or control monoclonal antibodies SS-1, SS-4, SS-7 and SS-8 (Center for Genetic Engineering and Biotechnology, Sancti Spiritus, Cuba), were added and incubated at 37 °C for 2 h in 0.2 % skim milk, 0.05 % Tween 20 in PBS, followed by six washes with PBS containing 0.05 % Tween 20. Bound antibodies were detected with horseradish peroxidase-conjugated goat anti-mouse IgG (SIGMA, USA), anti-Rat IgG (SIGMA, USA) anti-human IgG (1:10000, Jackson, USA) at 37 °C for 1 h, followed by washes. The reaction was detected after the addition of 3,3-5,5-tetramethylbenzidine and quantified using a microplate reader at 450 nm (BMG Labtech, Germany). This assay was also used for the evaluation of RBD antigenicity using sera from immunized animals, and from subjects that received Pfizer-BioNTech or Sputnik V vaccines, or are COVID-19 convalescents.

#### RBD to ACE2 plate-based binding assay

A competitive ELISA was performed to determine the inhibitory activity of the anti-RBD polyclonal sera on the binding of the hFc-ACE2 coated plates to an hFc-RBD-HRP conjugate. Briefly, the wells of ELISA plates were coated with 0.25 µg of recombinant hFc-AEC2 as described above. A mixture containing an hFc-RBD-HRP conjugate and serial dilutions of the sera were pre-incubated for 1h at 37 °C. A hundred microliters of the mixture were added to hFc-ACE2 coated plates and further incubated for 90 min at 37 °C. The binding of the HRP tagged RBD to the receptor was detected after the addition of 3,3-5,5-tetramethylbenzidine and reading at 450 nm. A similar assay was used to characterize the ability of the C-RBD-H6 PP and C-RBD-H6 HEK proteins to block the interaction of hFc-RBD-HRP with coated hFc-ACE2.

### 2.10. Microneutralization of live SARS-CoV-2 virus in Vero E6

The neutralization antibody titers were detected by a traditional virus microneutralization assay (MN50) using SARS-CoV-2 (CUT2010-2025/Cuba/2020 strain). Vero E6 cells (2×10^4^ per well) were seeded in 96-well plates one night before use. Animals’ sera were inactivated at 56 °C for 30 min. The samples were prepared by two-fold serial sera dilutions in the Eagle’s Minimal Essential Medium (MEM, Gibco, UK) containing 2 % (v/v) fetal bovine serum (Capricorn, Germany). SARS-CoV-2 strain at 100 TCID50 was incubated in the absence or presence of diluted sera for 1 h at 37 °C. Afterward, Vero E6 cell were overlaid with virus suspensions. At 96 h post-infection, the cells were inspected for signs of cytopathogenic effects (CPE) by optical microscopy and stained with neutral red (Sigma, USA). After three washes neutral red was dissolved in lysis solution (50 % ethanol, 1 % acetic acid) for 15 min at 25 °C, and optical density (OD) was detected at 540 nm. The highest serum dilution showing an OD value greater than the cut-off was considered as the neutralization titer. The cut-off value is calculated as the average of the OD of the cell control wells divided by two. The viral neutralizing titers (VNT50) were calculated as the highest serum dilution at which 50 % of the cells remain intact according to neutral red incorporation in the control wells (no virus added).

### 2.11. Cellular immune response

Long-term cellular immune response was evaluated in BALB/c mice using subcutaneous administration of a formulation containing an equal antigen to alum ratio. Animals receive 25 µg of the C-RBD-H6 PP antigen by the subcutaneous route in a 100 µL volume in a 0-14-35 days schedule. Blood samples were evaluated two weeks after the last immunization and animals were euthanized 3 months later to assess for systemic and lung resident cells response to *in vitro* antigen recall. Splenocytes were flushed out by perfusion using gentamycin supplemented PBS and lung resident lymphocytes were extracted after elimination of remnant blood by left auricular perfusion with PBS. To detach leukocytes from lung tissue, a lung dissociation enzyme mix (130-095-927, Miltenyi, Germany) was used according to manufacturer instructions and using a cell dissociator (GentleMACS Octo Dissociator, Miltenyi). CD3+ cells were further selected from lung suspension using negative selection with magnetic beads (130-095-130, Miltenyi) and both lung CD3+ cells and splenocytes live cells were counted using a flow cytometer (CyFlow, Sysmex, Germany).

For the re-stimulation assays, splenocyte or lung selected CD3+ suspensions were diluted to 10×10^6^ CD3+ live cells/mL and 50 µL of each sample was seeded in two 96-well U bottom tissue culture plates. Cells were re-stimulated with 50 µL of 20 µg/mL C-RBD-H6 PP, or just media (unstimulated) for 72 h, and the supernatant was analyzed at 1:2 dilution using Biolegend Deluxe cytokine kits for IL-2 (431004), IFNγ (430804), IL-6 (431304), IL-4 (431104) and TNFα (430204) following manufacturer’s instructions. Cells were transferred at that point to anti IFNγ coated ELISpot plates (Mabtech, Germany) and results were analyzed 24 h later according to established procedures.

### 2.12. ELISpot assay with samples from previous naturally infected individuals

PBMCs from subjects with previous natural infection with SARS-CoV-2 were isolated from 7 mL of whole blood collected using CPT tubes (Becton Dickinson, US), and store in liquid nitrogen until analyzed. After overnight resting the cells in Optmizer media (Gibco, Invitrogen, US) CD3+ live cells were counted using flow cytometry and seeded on activation plates at 5×10^4^ cells per well with 10 µg/mL of the recombinant C-RBD-H6 PP protein for 72 h. After transferring the cells to anti-IFNγ pre-coated plates (Mabtech, GE). The amounts of T cells secreting the cytokine were detected after 20 h of incubation as recommended by the manufacturer. All individuals gave their writing informed consent for the use of their samples.

### 2.13. Statistical analysis

Prism 8.4.3 software was used to generate dose-response curves for ELISA test and to calculate EC50 values when needed. One way-ANOVA with Sidak’s multiple comparisons tests-was used to determine the significance of differences and Spearman test was used to assess for parameters correlations. Wilcoxon matched paired test was used to perform paired analyzes for both stimulated/non-stimulated samples, the evolution of RBD specific titers and inhibition of ACE2 binding.

## 3. Results

### 3.1. Design of the C-RBD-H6 expression cassette

Recombinant C-RBD-H6 PP protein was designed as a potential subunit vaccine candidate against SARS-CoV-2 displaying e modular structure consists of: a) a globular central – RBD-domain comprising residues N_331_-K_529_ of the spike protein, b) additional N- and C-terminal segments including polar and flexible linkers rich in Glycine and Serine (Gly^9^-Ser^15^ and Gly^215^-Ser^229^). The N- and C-terminal extensions are aimed to modulate potential protein-protein interactions and facilitate protein purification through a high accessible hexa-Histidine tag (His^232^-His^237^). Topologically, the extensions are located in the opposite site of the protein respect to the receptor binding motif and its presence should sterically hinder potential aggregation problems associated to the presence of the exposed and disulfide bonded Cys^76^ and Cys^210^. (The C-RBD-H6 PP protein sequence is included as a supplemental material S1).

### 3.2. Expression and purification of the C-RBD-H6 PP

A construct for the expression in *P. pastoris* of the RBD from SARS-CoV-2 under control of the AOX1 promoter, denominated pPICZα-CtagRBDH6, was prepared as described in Materials and Methods and used to obtain RBD-expressing yeast clones. Fig. 1 (lanes A-C) shows the result of the analysis of culture supernatants from 3 randomly selected clones after induction with methanol. There is a noticeable smear in the supernatant from all three clones, ranging between 25 kDa and 50 kDa, that is absent from the control transformed with the empty vector (lane V, Fig. 1), and resolves into a ladder when probed by Western blotting with an anti-His6 antibody. Considering the presence of three-potential N-glycosylation sites in the sequence of C-RBD-H6 PP protein; and also that its molecular mass calculated from its cDNA sequence is 26 kDa the presence of a broad and diffuse band detected on SDS-PAGE analysis suggest that the C-RBD-H6 PP is being secreted in all three cases as a N-glycosylated protein. Based on signal intensity in Western blotting, clone C was denominated X33-23 and selected for further work (Fig. 1). The C-RBD-H6 PP protein was purified from the supernatant following the procedures described in Materials and Methods, by IMAC, followed by polishing using a RP to obtain a final preparation with high purity. After purification, the C-RBD-H6 PP was obtained with a final yield between 30–40 mg/L of culture medium, with more than 98% of purity at the end of the downstream process (Fig. 2 and 3).

**Fig. 1.**
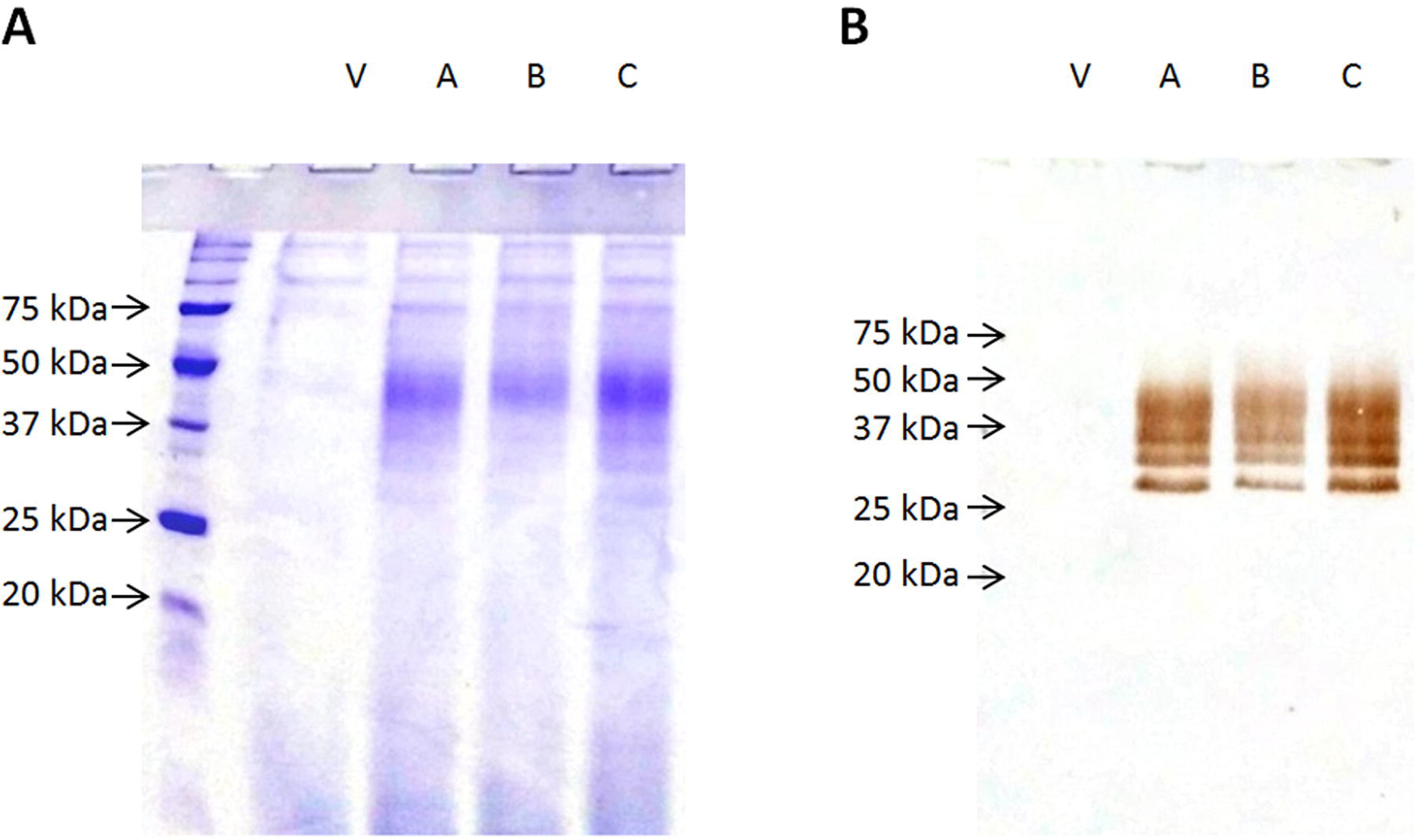
Analysis of culture supernatants from randomly selected RBD-secreting *P. pastoris* clones. In all cases 10 µL of each sample were analyzed under reducing conditions. Panel A: Coomassie Blue-stained 12.5 % SDS-PAGE; panel B: Western blotting of an identical gel with an anti-His tail antibody. Lane V: Strain transformed with the empty vector. Lanes A, B and C: clones transformed with the RBD plasmid.

**Fig. 2.**
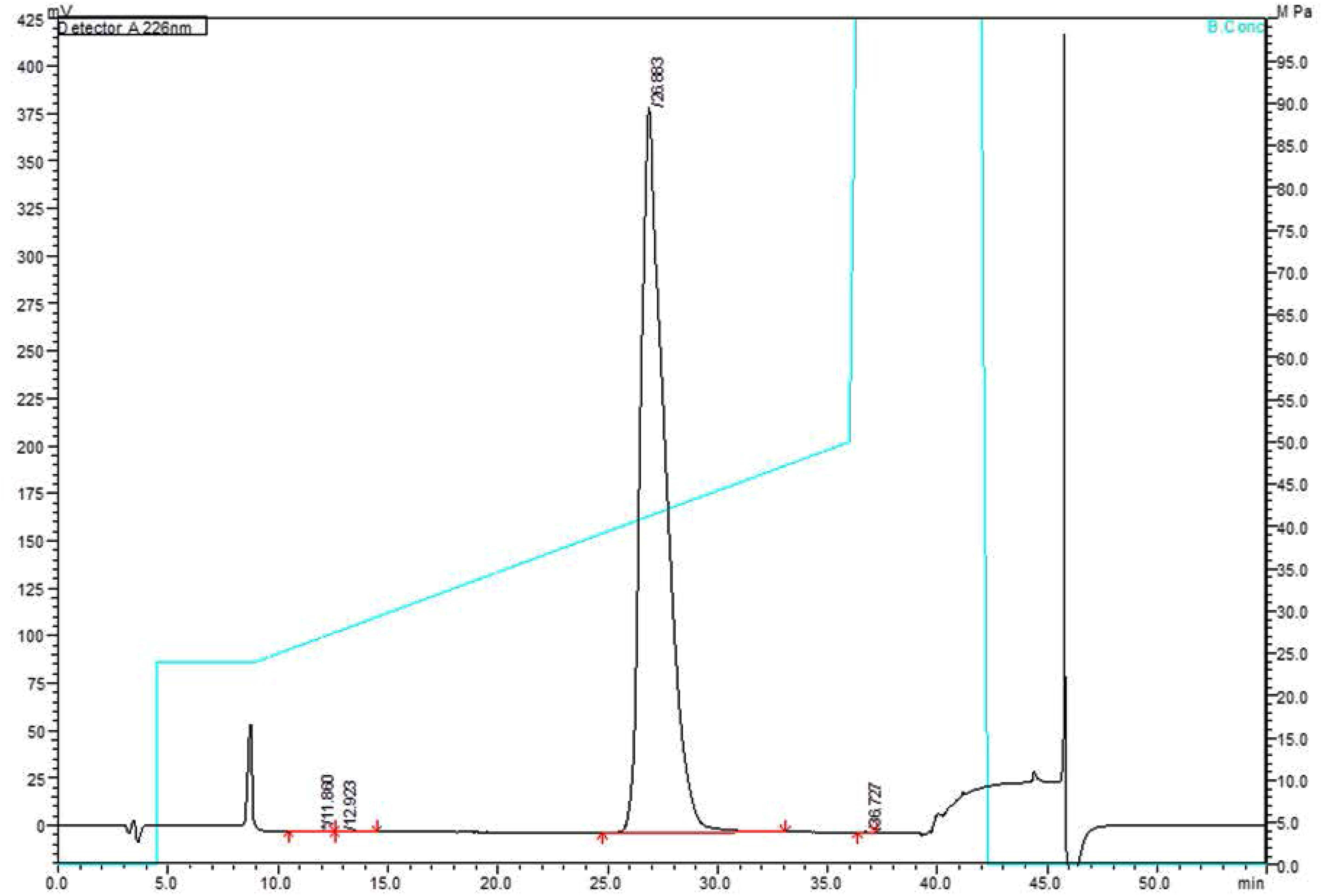
RP-HPLC. Analysis of C-RBD-H6 PP protein produced in *Pichia pastoris*, with 98.6 % of purity; analyzed on a reversed phase C8 Vydac analytical column. The gradient is shown with a blue line.

**Fig. 3.**
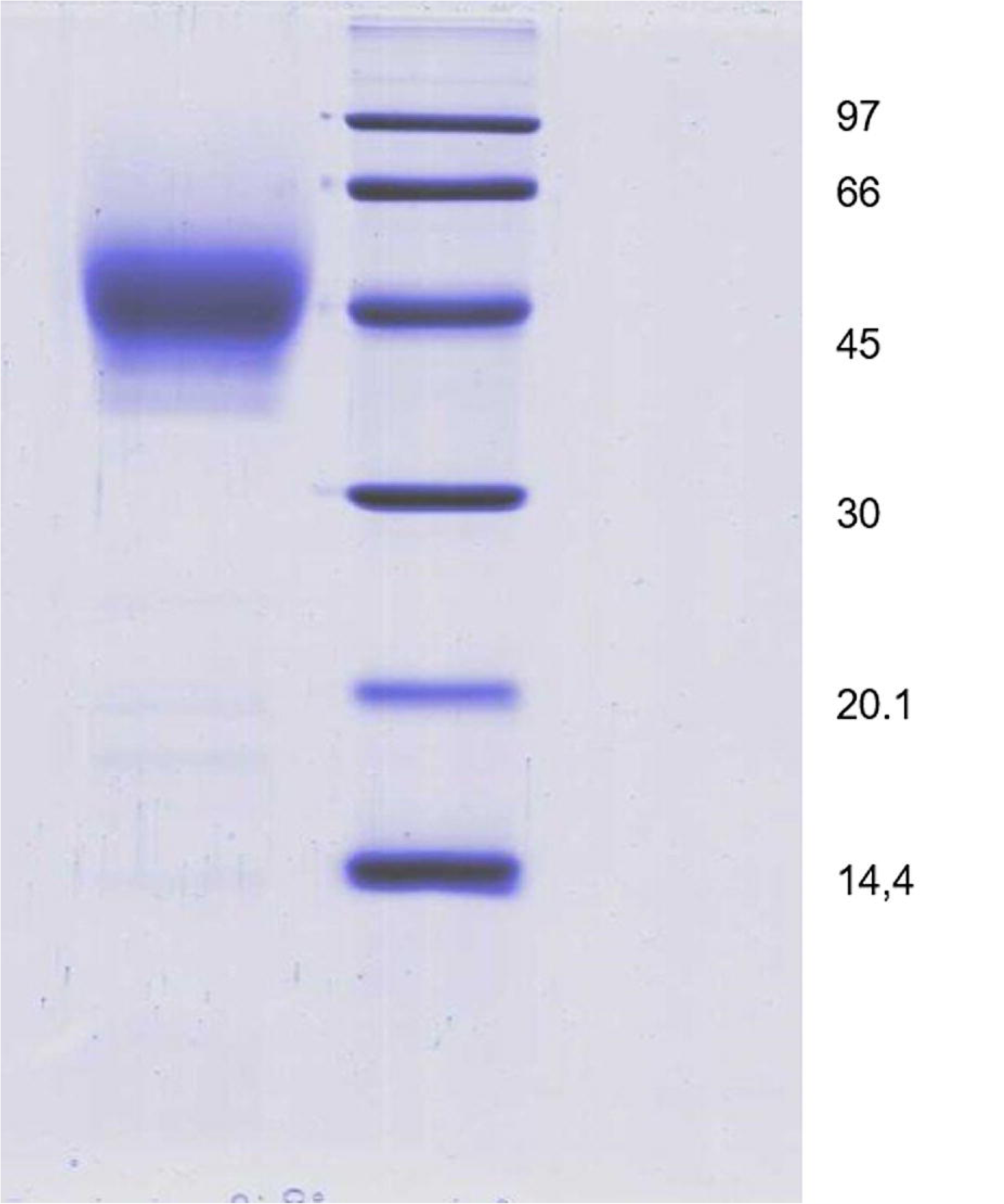
Protein electrophoresis. Coomassie Blue stained 12,5 % SDS-PAGE gel of 10 μg of the purified C-RBD-H6 PP under reducing conditions. Lane 1: C-RBD-H6 PP protein; Lane 2 Molecular weight markers.

### 3.3. ESI-MS analysis for determining the accurate molecular mass, the verification of the amino sequence and the assessment of disulfide bonds of the deglycosylated C-RBD-H6 PP protein

The C-RBD-H6 PP protein deglycosylated with PNGase-F was analyzed by ESI-MS and the multiply charged spectrum is shown in Fig. 4.A. Deconvolution of this ESI-MS spectrum (Fig. 4.B) showed a very intense signal at 26069.54 Da. This molecular mass agrees very well with the calculated molecular mass (26069.87 Da) for the C-RBD-H6 PP considering the presence of four intramolecular disulfide bonds and the two out the three potential N-glycosylation sites transformed into Asp residues due to the action of PNGase-F. Also a low-abundance signal (see star in Fig. 4.B) indicates that a minor fraction of this molecule is devoid of the extension of four N-terminal amino acids (- NSWF) not belonging to the RBD protein.

**Fig. 4.**
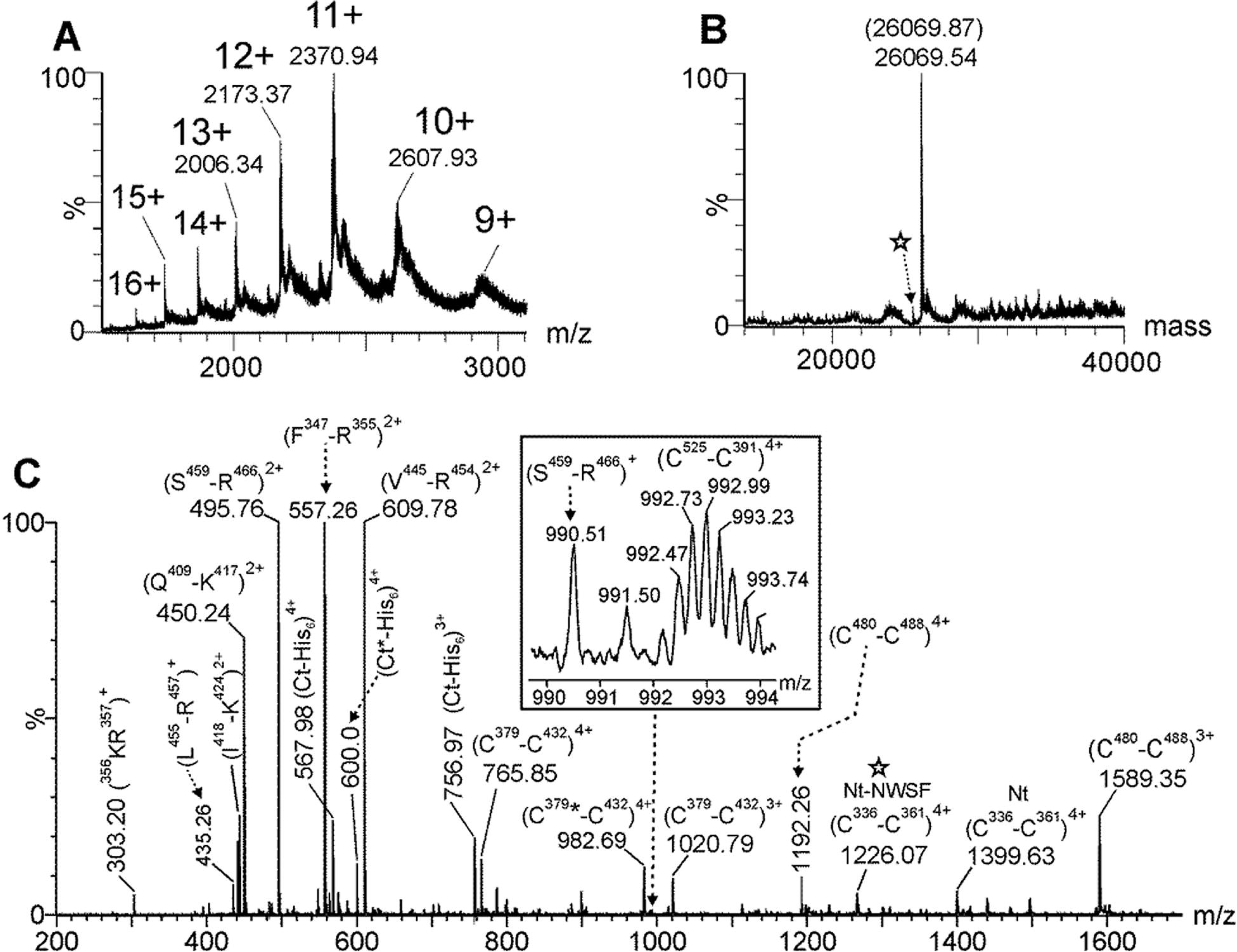
ESI-MS spectra. (A) Multiply-charged and (B) deconvoluted ESI-MS spectra of C-RBD-H6 PP protein previously N-deglycosylated with PNGase-F. The parenthesis in (B) shows the expected molecular mass considering the presence of four intramolecular disulfide bonds and the Asn^331^ and Asn^343^ transformed into Asp residues by the PNGase F during the deglycosylation step. (C) ESI-MS spectrum of the tryptic peptides generated by an in-solution buffer-free trypsin digestion protocol of the N-deglycosylated C-RBD-H6 PP protein. The star in (B) and (C) indicates the low-abundance species that do not contain the four residues (-NWSF) located at the N-terminal end. Nt and Ct-His_6_ represents the N-terminal end and the His_6_-tag C-terminal end peptides of C-RBD-H6 PP protein, respectively.

To verify the amino acid sequence as well as the disulfide bonds the deglycosylated protein was digested with trypsin using an in-solution buffer-free digestion protocol developed in our group [16;20]. ESI-MS analysis of the tryptic peptides is shown in Fig. 4.C. Full-sequence coverage of C-RBD-H6 PP was verified and the assignment for signals observed in this mass spectrum is summarized in Table 1. ESI-MS/MS analysis of the signal detected at *m/z* 1399.64, 4+ confirmed that Asn^331^ and Asn^343^ (two out the three potential N-glycosylation sites) were transformed into Asp residues by the action of PNGase F (see underlined residues in the Table). The four disulfide bonds (C_336_-C_361_, C_379_-C_432_, C_391_-C_525_ and C_480_-C_488_) present in the native S protein of SARS-CoV-2 were also detected in this ESI-MS spectrum (Fig. 4.C and Table 1). Tryptic peptides containing free cysteine residues and S-S scrambling variants were not detected.

**Table 1.**
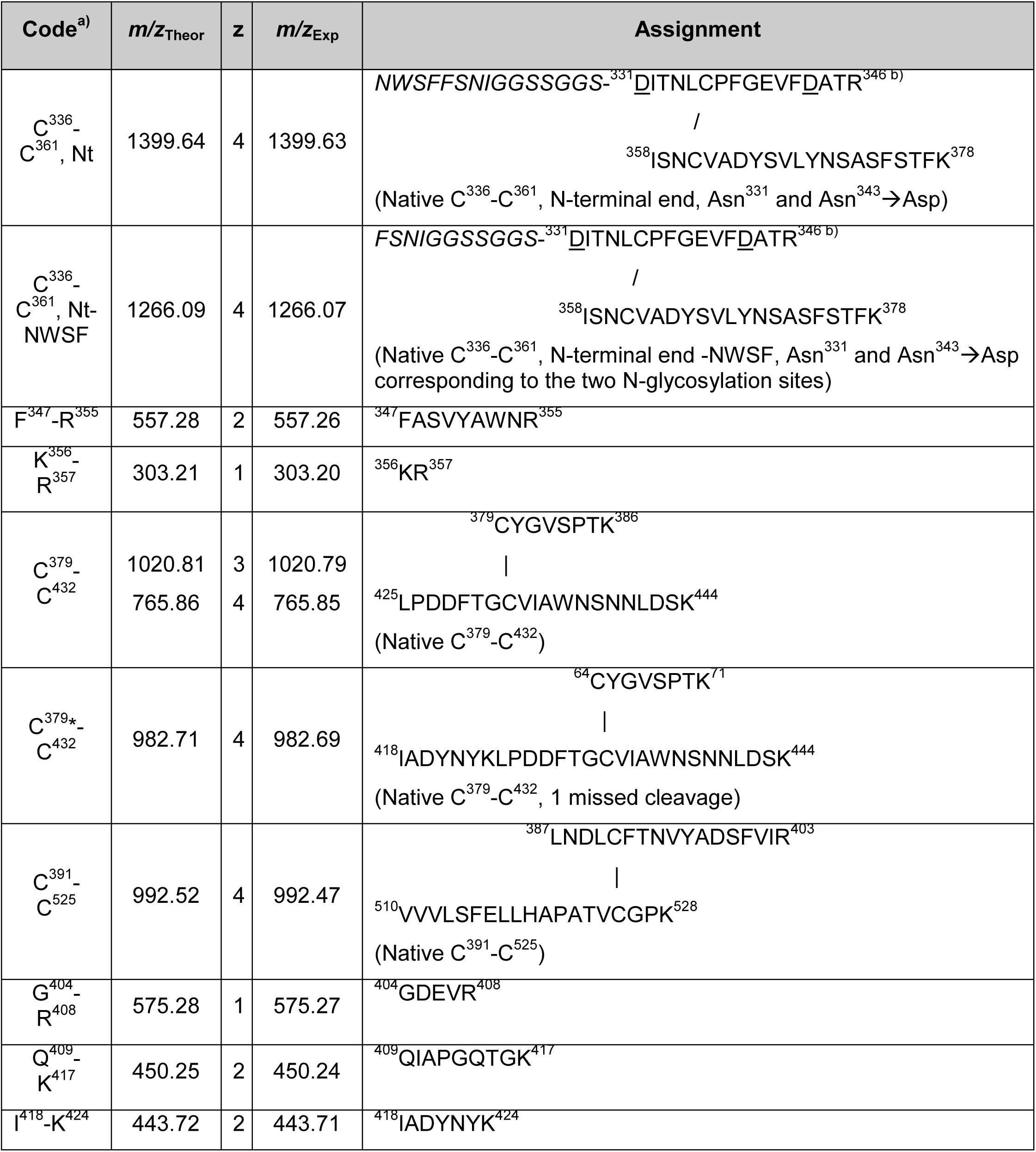
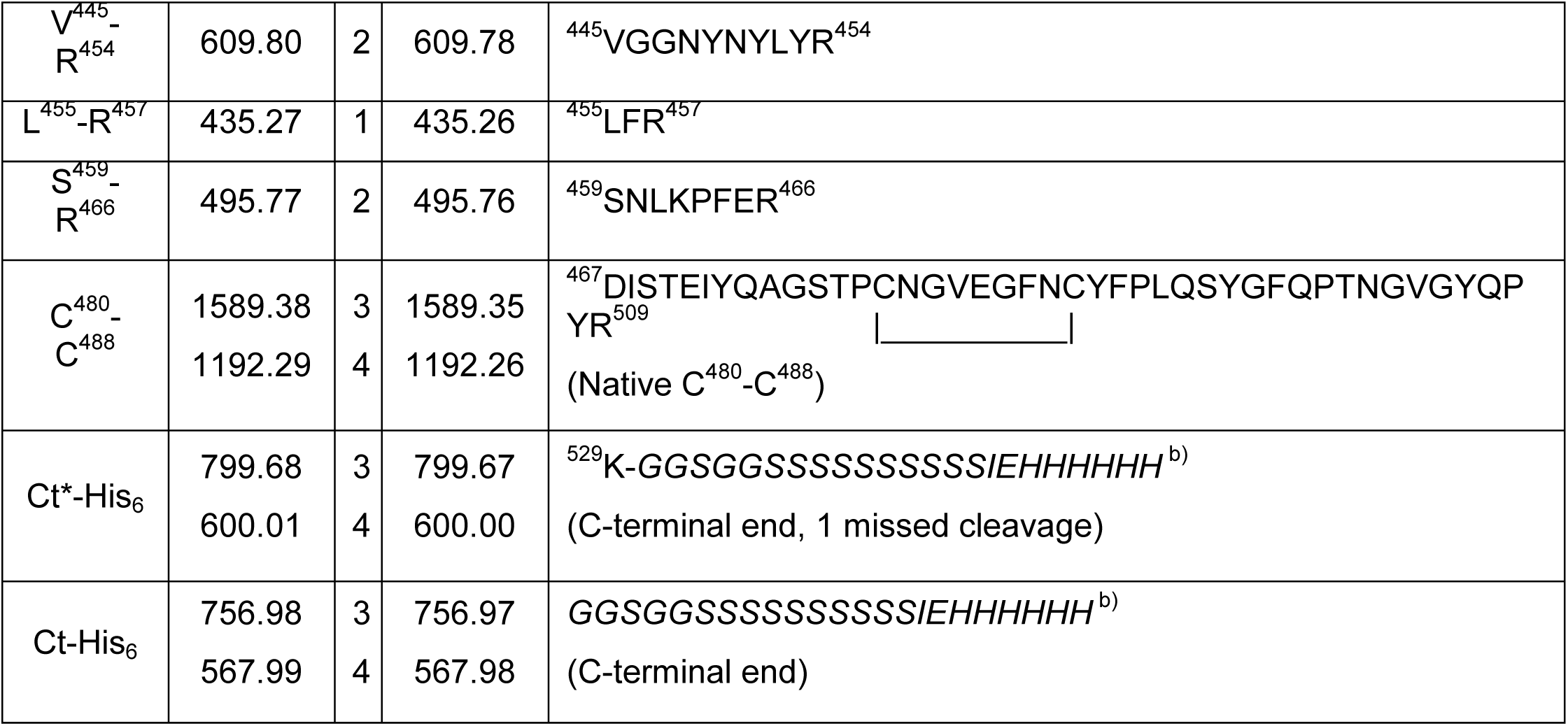
Summary for the sequence verification of the N-deglycosylated C-RBD-H6 PP considering the ESI-MS analysis of tryptic peptides generated by the in-solution buffer-free digestion. Nt: N-terminal end, Ct-His_6_: His-tag C-terminal end. C^#^-C^#^ corresponds to tryptic peptides linked either by intermolecular disulfide bonds or a tryptic peptide that contains an intramolecular disulfide bond in its structures. *m/z*_calc_ correspond to the calculated *m/z* values for all tryptic peptides generated by the in-solution buffer-free digestion of the N-deglycosylated protein. *m/z*_exp_ correspond to the experimental *m/z* values for all tryptic peptides observed in the ESI-MS analysis shown in Fig. 4.C. Regions of the sequence written in italic do not correspond to the RBD of SRAS-CoV-2 and were inserted in the cloning stage, while underlined residues indicate the conversion of N-glycosylated asparagines (Asn^331^ and Asn^343^) into aspartic acid residues by the action of PNGase-F (Asn^331^ and Asn^343^→Asp).

ESI-MS analysis of the deglycosylated C-RBD-H6 PP protein and its derived tryptic peptides confirmed that N-glycans in its structure increased considerably its molecular mass by SDS-PAGE analysis.

### 3.4. Surface Plasmon Resonance characterization of the RBD-ACE2 binding affinity

As shown in Fig. 5, there was no appreciable signal in terms of response units (RU) when measurements were performed a non-related protein used as negative control for BIACORE experiments, immobilizing mFc-ACE2 ligand by Fc region on to a Prot A chip. In contrast, the association rate for C-RBD-H6 PP protein to mFc-ACE2 was 5.4×10^5^ M^−1^·s^−1^, with a dissociation rate of 7.7×10^−3^ s^−1^. The equilibrium was reached between 25 and 30 seconds, with an estimated dissociation constant of K_D_ = 14.3×10^−9^ M. The association/dissociation rates as well as K_D_ were in the expected ranges, according to the previously reported results for the RBD-ACE2 molecular interaction [25].

**Fig. 5.**
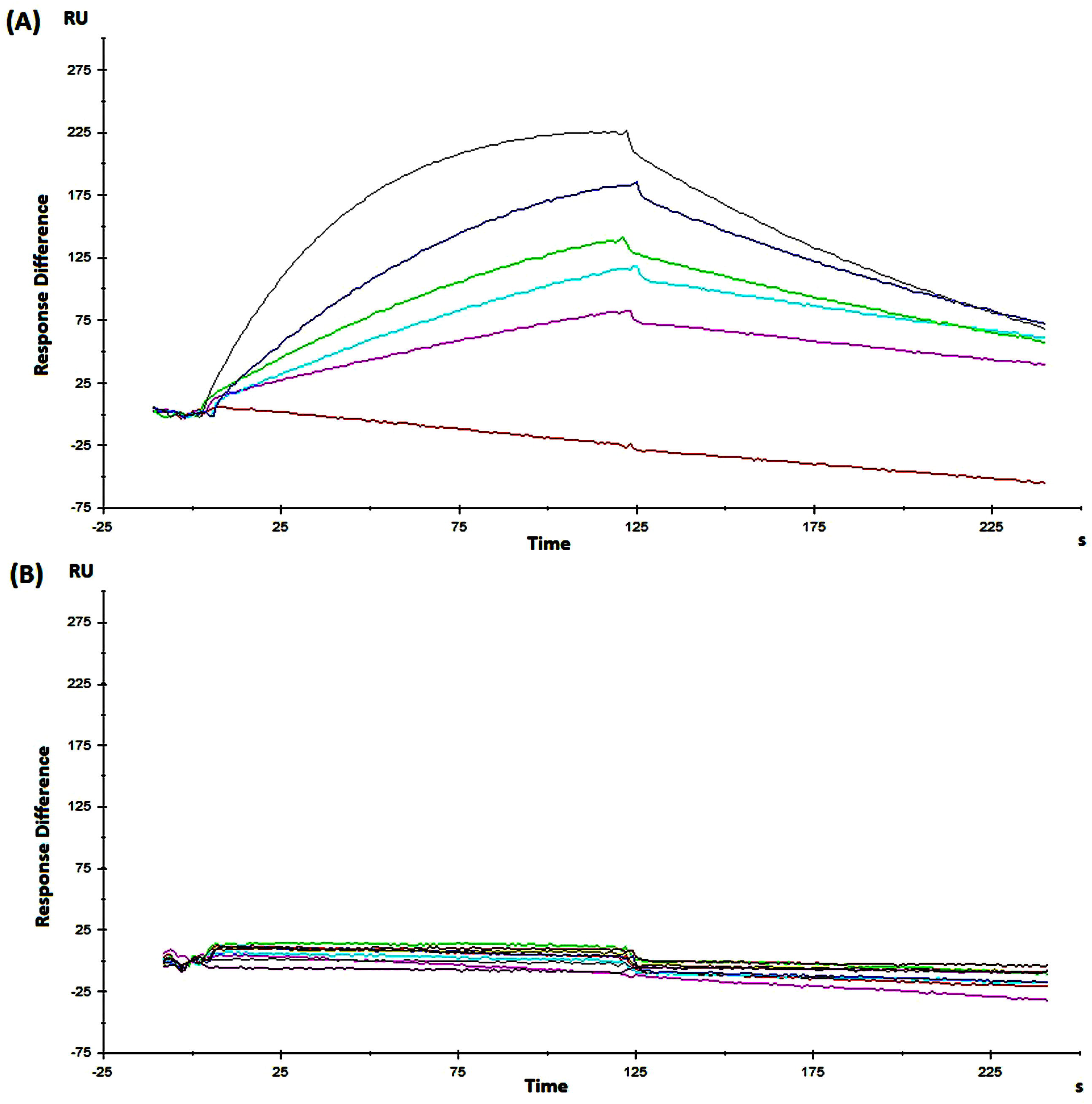
SPR analysis from C-RBD-H6 PP protein obtained in *P. pastoris* interacting with mFc-ACE2 receptor in a single-cycle BIACORE experiment. (A) Sensorgrams corresponding to one of the replicates of the protein dissolved in PBS, pH = 7.2, (B) Non-related protein (also expressed in yeast), used as negative control for the interaction with immobilized mFc-ACE2 cell receptor. As expected, there were no significant signals for the protein used as negative control; compared to the curves obtained for C-RBD-H6 PP protein in the same experimental conditions (similar axis scale is shown).

### 3.5. Secondary structure analysis by CD Spectroscopy

Fig. 6 shows the far UV CD spectrum of the C-RBD-H6 PP protein revealing characteristic bands similar to the observed in other recombinant RBD proteins reported previously [8], with maxima at 192 and 231 nm – due to the aromatic contribution - and minimum at 207 nm. Furthermore, as shown in Table 2 the secondary structure content of the protein estimated by BeStSel (7.9% helix, 28.7% beta - antiparallel, relaxed and right-handed-, 12.9% turn y 50.5% others) is very similar to the values assigned using the 3D coordinates [11]. Moreover, as observed in Fig. 7 the near UV CD spectrum of the protein is well structured, with bands at 263, 269, 277, 281 and 299 nm, indicative of the presence of well-packed aromatic and cysteine residues as expected for a properly folded protein.

**Fig. 6.**
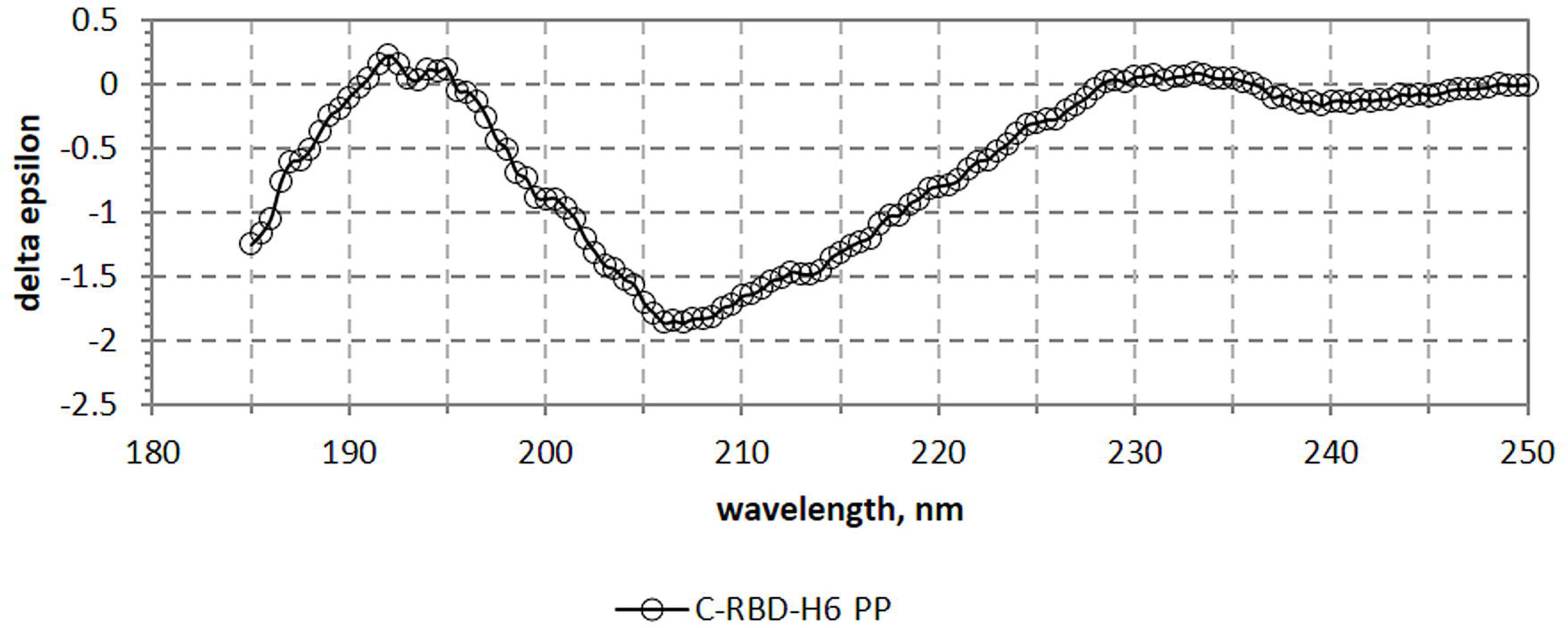
Far UV CD spectrum of the C-RBD-H6 PP protein.

**Fig. 7.**
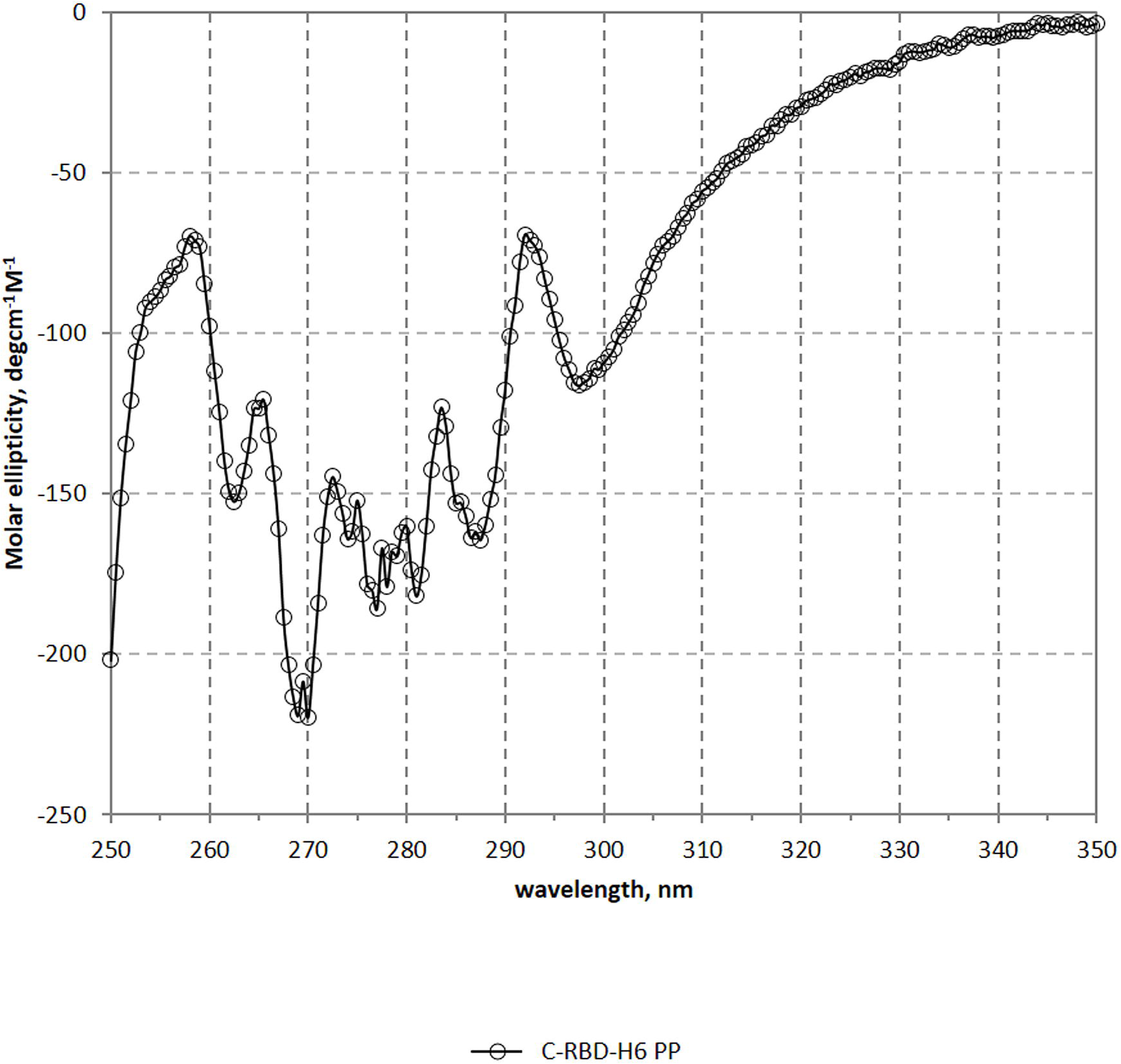
Near UV CD spectrum of the C-RBD-H6 PP protein. Bands at 263, 269, 277, 281 and 299 nm, indicate the presence of well packed aromatic and cystine residues.

**Table 2.** Secondary structure content of the protein estimated by CD (BeStSel) and 3D coordinates (DSSP)

| Secondary Structure Element | Secondary Structure Content, % |  |
| --- | --- | --- |
|  | Method |  |
|  | BeStSel | DSSP |
| Helix | 7.9 | 9.3 |
| Beta antiparallel | 28.7 | 22.4 |
| Beta parallel | 0.0 | 0.0 |
| Turn | 12.9 | 22.4 |
| others | 50.5 | 45.9 |
| Helix1 (regular) | 3.3 | - |
| Helix2 (distorted) | 4.6 | - |
| Beta Antiparallel_1 (left-handed) | 0.9 | - |
| Beta Antiparallel_2 (relaxed) | 12.0 | - |
| Beta Antiparallel_3 (right-handed) | 15.8 | - |

### 3.6. Antigenicity analysis of C-RBD-H6 PP

To confirm the antigenicity of the yeast-derived protein, several ELISA tests were performed with SS-1, SS-4, SS-7 and SS-8 anti-RBD monoclonal antibodies obtained immunizing with RBD-H6 HEK protein produced in the mammalian cells HEK293T. One of these monoclonal antibodies (SS-8) inhibits the protein binding to the ACE2 receptor with an IC50 of 60 ng/mL. The binding to C-RBD-H6 PP protein was compared to the binding to C-RBD-H6 HEK as shown in Fig. 8.A,E. The reactivity does not differ between the two proteins. Human sera with high neutralization titers for live SARS-CoV-2 viral challenge of Vero E6 cells from eight COVID-19 naturally infected patients and seven individuals that already received Pfizer-BioNTech or Sputnik V complete vaccine schedules were incorporated into the testing panels. None properly folded C-RBD-H6 protein produced as inclusion body in BL-21 *E coli* strain was used as a negative control. C-RBD-H6 PP protein obtained from yeast was able to maintain antigenicity comparable to the protein purified from mammalian cells (Fig. 8.B,F and C,G). Minimal signals were detected with the sera from animals immunized with RBD protein expressed in *E. coli*, corroborating the need for a correctly folded protein to ensure the exposition of immunodominant epitopes (data not shown). Further characterization was conducted using polyclonal sera from mice and NHP immunized with C-RBD-H6 PP and C-RBD-H6 HEK proteins, and with known capacity to neutralize SARS-CoV-2 infection. All polyclonal sera similarly detect the RBD protein expressed in both heterologous expression system (Fig. 8.D,H).

**Fig. 8.**
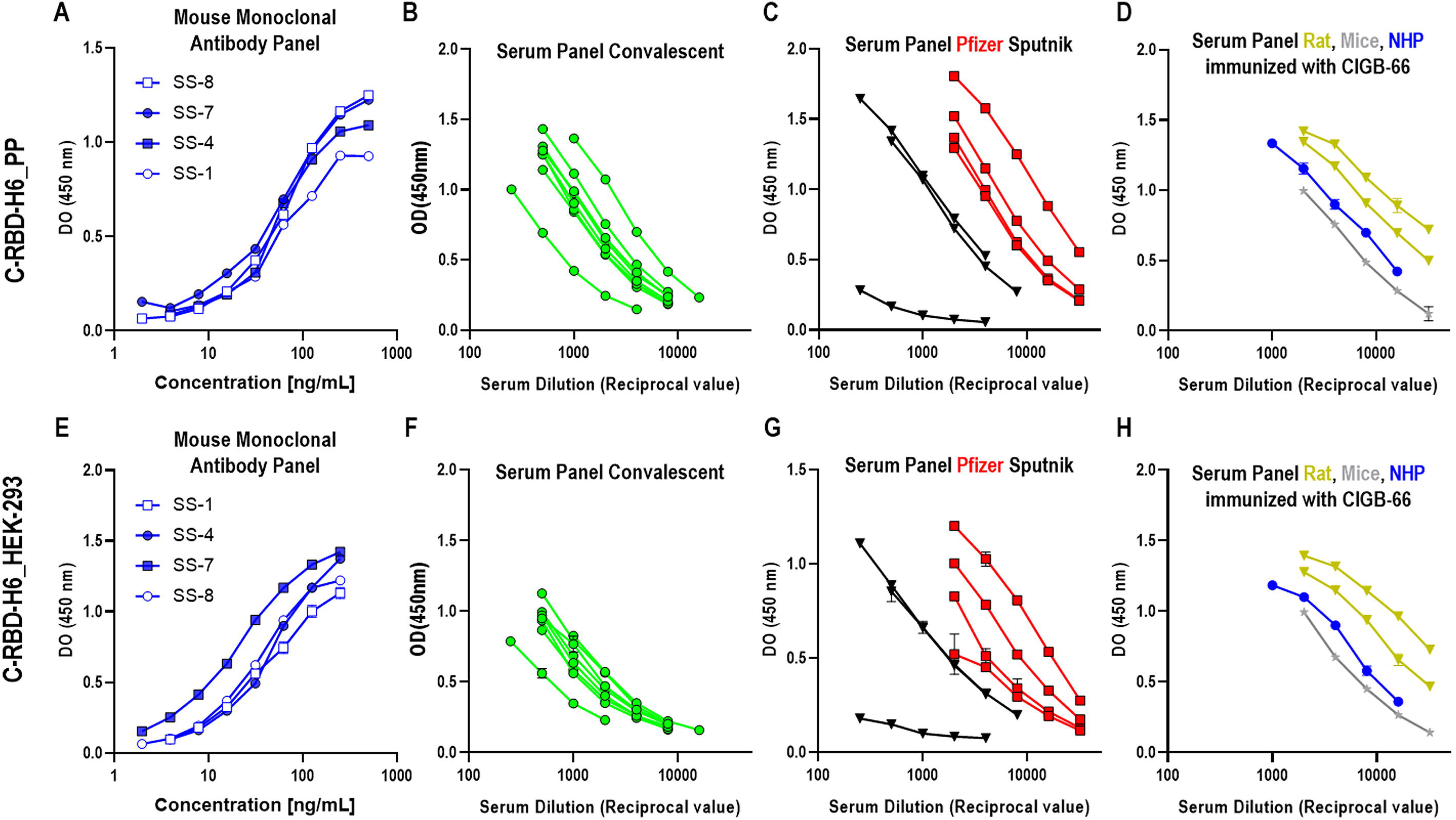
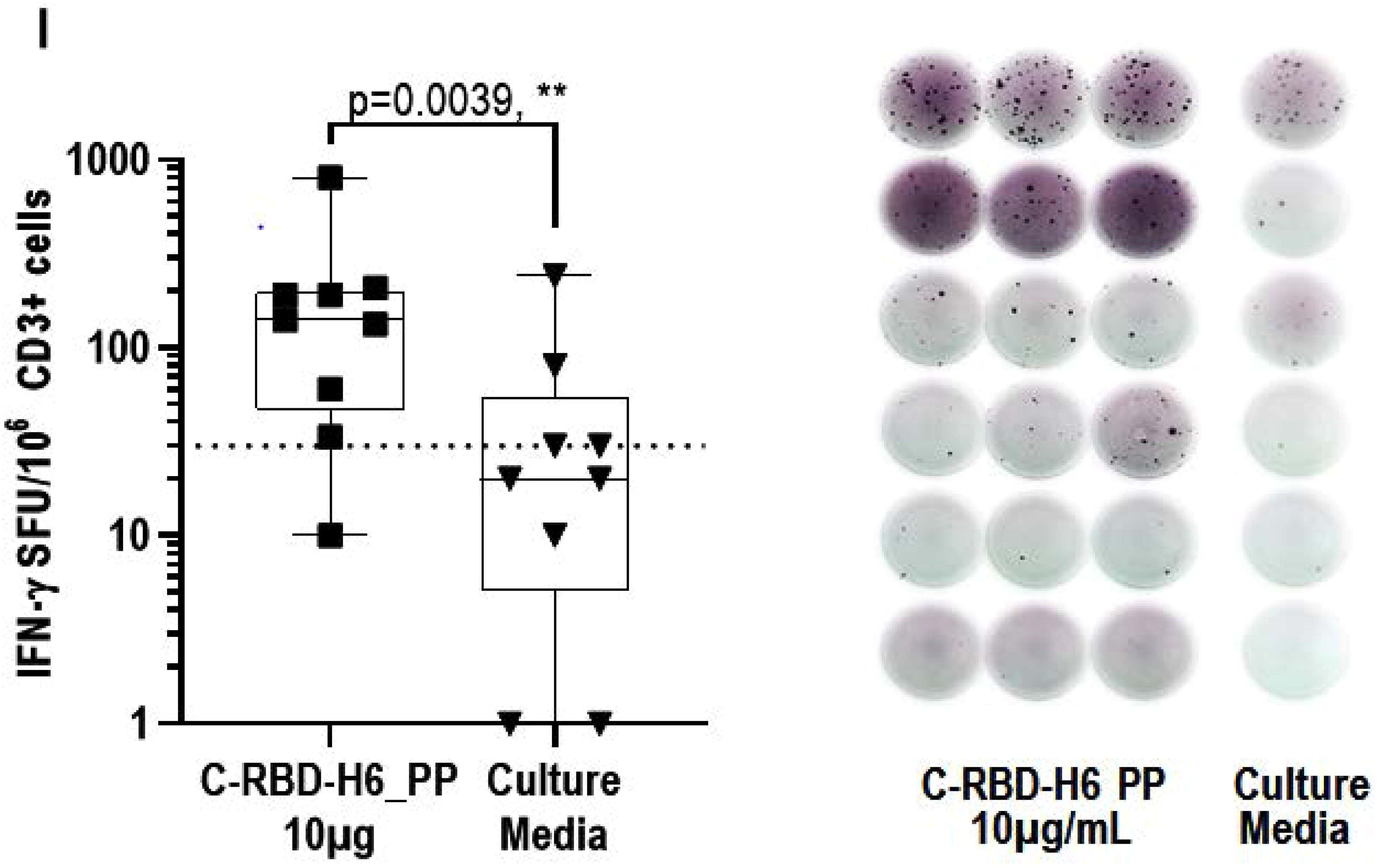
Antigenicity of the RBD protein produced in *P. pastoris* yeast. C-RBD-H6 PP (upper panel) or RBD-H6 HEK (lower panel) proteins were used for coating ELISA plates and for animal immunization. Sera and monoclonal antibodies were used in serial two-fold dilutions. (A,E). SS-1, SS4, SS-7 and SS-8 monoclonal antibodies; (B,F). COVID-19 sera from convalescents; (C,G) Sera from subjects immunized with Pfizer-BioNTech (red squares) and Sputnik vaccines (black triangles); (D,H). Mice (grey) and NHP (blue) sera; (**I**). C-RBD-H6 PP stimulates INFγ secretion in CD3+ cells from naturally infected individuals. Wilcoxon matched paired test.

The C-RBD-H6 PP protein was also used to recall *in vitro* a cellular response in terms of IFNγ secretion using ELISpot from PBMCs of COVID-19 naturally infected subjects with at least three months from hospital release. As shown in Fig. 8.I stimulation with 10 µg/mL of the yeast-produced protein induce IFNγ secretion from COVID-19 naturally infected subjects PBMCs.

### 3.7. Functionality of yeast-expressed RBD protein

#### ACE2 receptor binding competition assay

The functionality of the recombinant protein C-RBD-H6 PP was confirmed by the assessment of its ability to bind to the chimeric hFc-ACE2 receptor bound to ELISA plates and also to bind to the ACE2 receptor in Vero E6 cells both in solution or attached to the plate. To evaluate the binding of the protein to hFcACE-2, a serial two-fold dilution curve of the test protein starting from 100 μg/mL was mixed with HEK293T produced hFc-RBD-HRP conjugate at a fixed concentration and added to hFc-ACE2 coated plates. As shown in Fig. 9.A, two independent C-RBD-H6 PP protein preparations effectively displaced the hFc-RBD HEK in the same magnitude that the soluble hFc-ACE2 and the recombinant RBD-H6 HEK protein obtained in HEK293T cells. Two non-related chimeric proteins P64K-VEGFH6 (expressed in *E. coli*), and hFc-VEGFR2 (produced in HEK293T cells) were used as negative controls

**Fig. 9.**
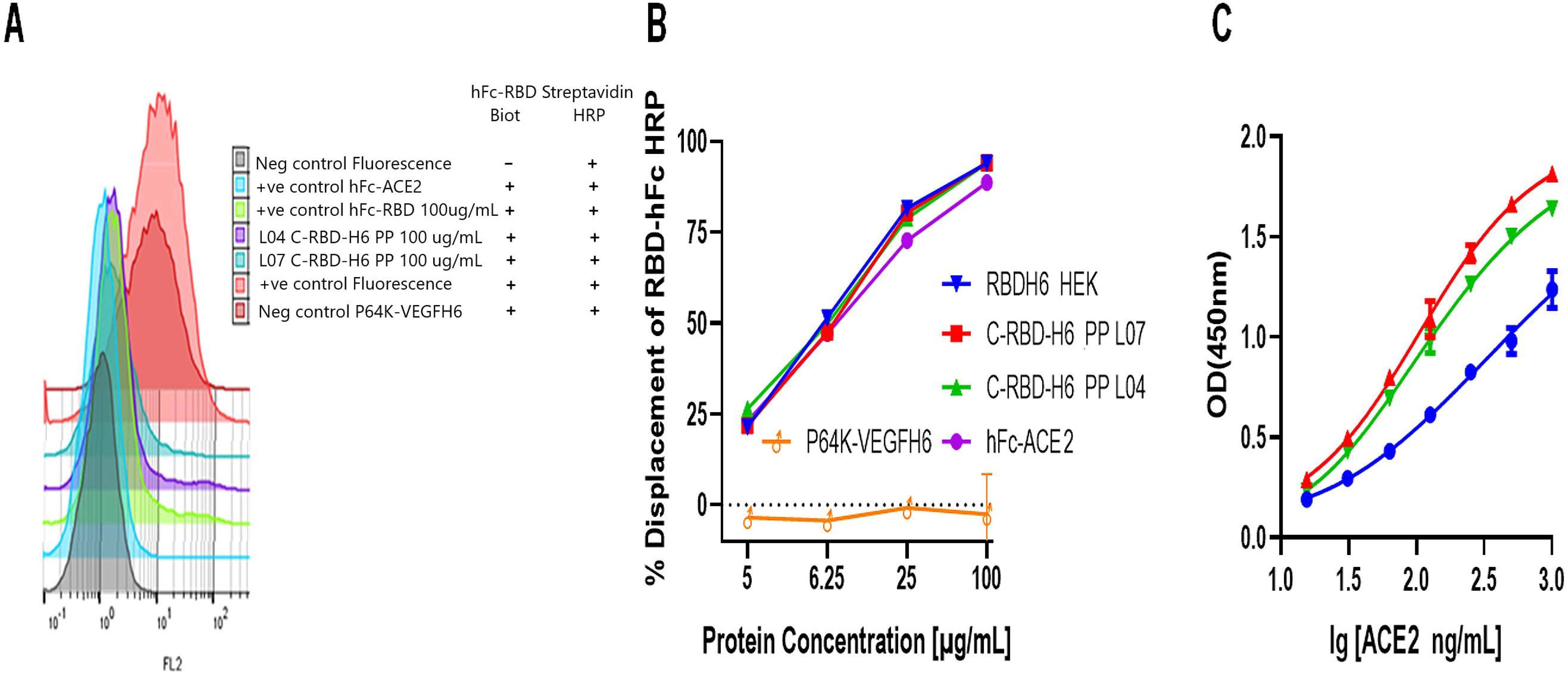
Binding inhibition capacity of two independent protein preparations of the C-RBD-H6 PP (L04 and L07). (A). The C-RBD-H6 PP protein displaced the hFc-RBD-HRP from the hFc-ACE2 coated ELISA plates, and (B) from the ACE2 receptor in Vero E6 cells. (C). The C-RBD-H6 PP protein was recognized by the soluble ACE2 receptor. P64K-VEGFH6 and hFc-VEGFR2 non-related chimeric proteins were used as negative controls.

A similar study was conducted on paraformaldehyde-fixed Vero E6 cells (Fig. 9.B) As expected, no displacement of biotinylated hFc-RBD from the cell membrane was observed in either case using as negative controls the non-related chimeric VEGF protein fused to the N-terminal of *Neisseria meningitidis* (P64K-VEGFH6) or the human Fc (VEGFR2-hFc), and 100 % of the signal was lost when the C-RBD-H6 PP protein, soluble hFc-ACE2 or mFc-ACE2 were added (Fig. 9.C). These results suggest that the yeast-expressed C-RBD-H6 PP protein can bind efficiently to the cell-surface ACE2 receptor.

Further characterization of ACE2 binding was conducted in solution using yeast and mammalian-derived proteins coated in different concentrations in ELISA plates. ACE2 EC-50 on RBD coated plates result in values ranging between 100 and 300 ng/mL in agreement with the values reported for commercial sources of RBD and with our data using RBD variants purified from HEK293T supernatant. Using the same system stability of the protein was verified by incubating the protein at 4 °C, 37 °C, and 50 °C for 2 h without significant loss in ACE2 binding activity (data not shown)

These results confirm the specific binding of the yeast-expressed RBD with SARS-CoV-2 receptor ACE2 either cell-bound or soluble, indicating that the antigen herein obtained is functional.

#### Yeast-expressed RBD protein elicited RBD-ACE2 receptor binding inhibition and SARS-CoV-2 neutralizing antibodies in rodents and NHP

To evaluate the immunogenicity of C-RBD-H6 PP protein, BALB/c mice were immunized thrice at 7 days intervals by the intraperitoneal route with 50µg of the protein. The IgG antibody response in mice at days 21 and 35 shows that the C-RBD-H6 PP protein was able to induce antibody responses in ELISA coated with the RBD protein produced in mammalian HEK293T cells. Seroconversion was achieved a week after the first boost (Fig. 10.A). The administration of a second booster at day 21 significantly increases both RBD-specific IgG titers and the inhibitory potential of the sera to reduce the protein binding to its cognate receptor ACE2 a fact that correlates with live SARS-CoV-2 neutralization (Fig. 10.B,C).

**Fig. 10.**
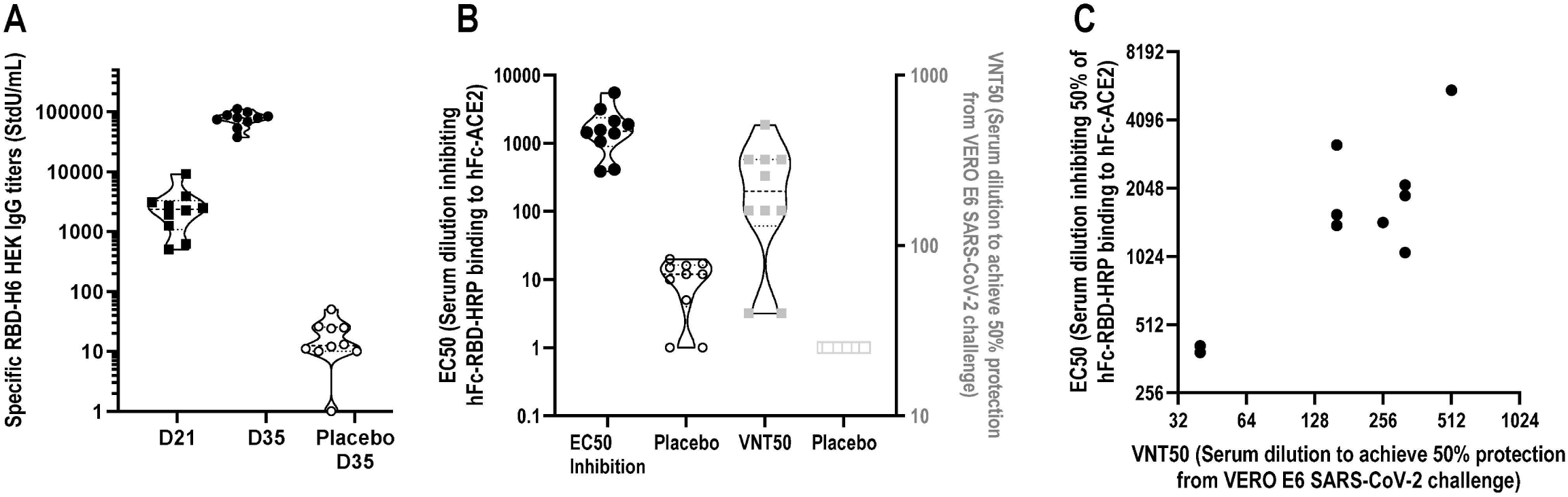
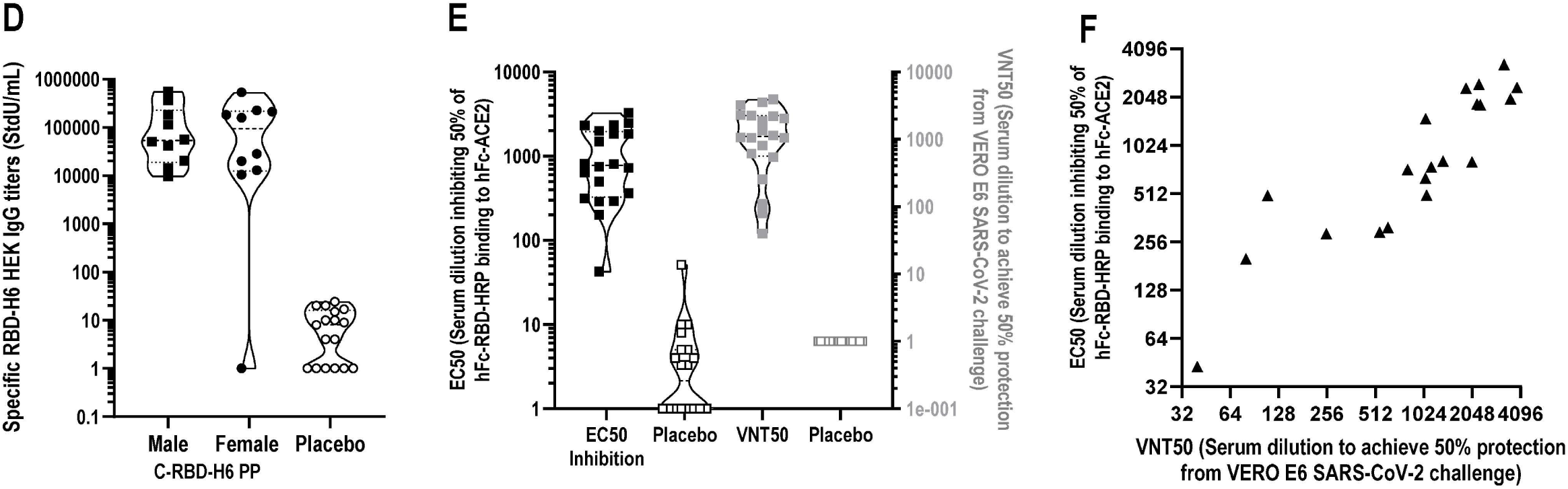
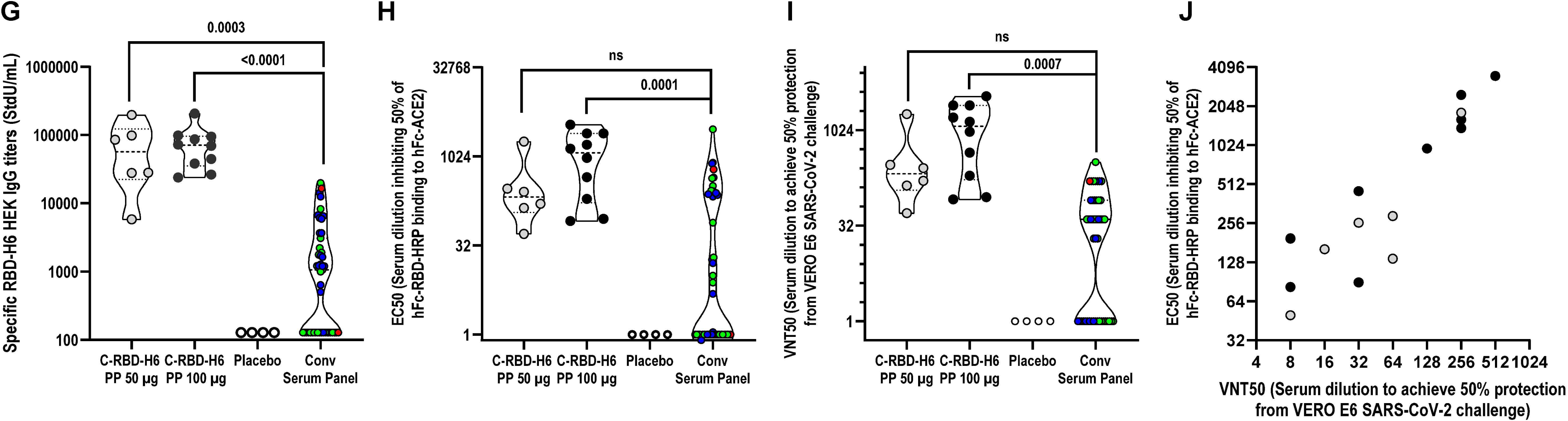
Immunogenicity of C-RBD-H6 PP protein in three animal species using different immunization routes and schedules. (A) Evaluation of RBD-specific IgG in BALB/c mice 7 days after second and third intra-peritoneal immunization (n=10); (B). Evaluation in BALB/c mice of EC50 for the ACE2 binding inhibition and PRNT in the microneutralization assay (n=10); C. Correlation analyses of the ACE2 binding inhibition and microneutralization tests in mice (C). Sperman, r=0.6482, p=0.0478); (D). Evaluation of RBD-specific IgG in SD rats 3 days after 10 weakly subcutaneous immunization (n=10); (E). Evaluation in SD rats of EC50 for the ACE2 binding inhibition and PRNT in the microneutralization assay (n=20); (F). Correlation analyses of the ACE2 binding inhibition and microneutralization tests in rats (Spearman, r=0.9233, p=< 0.0001); (G). Evaluation of RBD specific IgG in NHP with 50 µg and 100 µg dose 14 days after three subcutaneous immunizations every second week with 6 and 10 animals respectively; (H). Evaluation in NHP of EC50 for the ACE2 binding inhibition; (I). Evaluation in NHP of EC50 for the PRNT in the microneutralization assay. (J). Association Correlation analyses of the Inhibition and microneutralization tests in NHP (Spearman, r=0.8994, p<0.0001).

The C-RBD-H6 PP protein was also tested in SD rats using intramuscular administration of 9 µg every 7 days for 10 weeks. Assessment of IgG and RBD-ACE2 receptor binding inhibition three days after the last immunization indicates high RBD-specific antibody titers that correlate with the inhibitory titer. The former also display a high correlation with the neutralization titer of live SARS-CoV-2 virus in Vero E6 cells (Fig. 10.D,F).

The C-RBD-H6 PP protein evaluation in NHP using short intramuscular administration schedule on days 0-14-28, and two dose levels indicate a dose-response effect with seroconversion of 83 % (5 of 6 animals) and 100% (the 10 animals) for the 50 µg and 100 µg dose respectively after the first booster and a 100 % seroconversion in both schedules after the second booster. Total IgG titers increased to 44,240 StdU/mL and 62,435 StdU/mL respectively for monkeys included in the low and high-dose groups. In both cases, the titers were significantly higher than those detected in the convalescent panel of sera. The geometric median ACE2 binding inhibition titer for 50 µg dose was 1:230, while a significantly higher value of 1:705 was detected for the animals receiving the 100 µg dose. Both values were higher than those detected in a panel of COVID-19 convalescent subjects (p<0.001, Kruskal-Walis). The higher titers for the 100 µg dose correlate with and enhancement in the inhibition of RBD-ACE2 binding for this group. The analysis of the neutralization titer of live SARS-CoV-2 virus in Vero E6 cells corroborates these findings, indicating that 50µg dose is sufficient to induce ACE2 binding inhibition titers with an EC50 geometric mean of 1:66 serum dilution, a value 6 times higher than the reported for the convalescent panel. A significant increase to 1:141 in this parameter was detected for the 100 µg dose pointing to a dose-related effect (Fig. 10.G,J).

The specific IgG antibody and the inhibition of RBD-ACE2 binding were only detected in C-RBD-H6 PP protein-inoculated animals but not in control animals. The immune responses evidenced the boosting effect and the specific dose-dependent effect, as compare to negative results for control animals.

#### Cellular response

Cellular response recall was evaluated three months after the last immunization in BALB/c mice receiving subcutaneous doses of 25 µg in alum in a 0-14-35 days schedule. The evaluation of the presence of a memory response in the animal’s spleens indicates a significant induction of clones secreting IFNγ in response to incubation with the C-RBD-H6 PP antigen. Furthermore, the analyses of the supernatant of the recall reaction point to the predominant induction of IFNγ followed by IL-2, IL-6 and to a lower extend TNFα and IL-4 (Fig. 11). Our findings in the systemic compartment (Fig. 11.A) were similar to those found for CD3+ cells enriched from mice lungs (Fig. 11.B), indicating that the response can also be recalled in the organ primarily affected by SARS-CoV-2 infection.

**Fig. 11.**
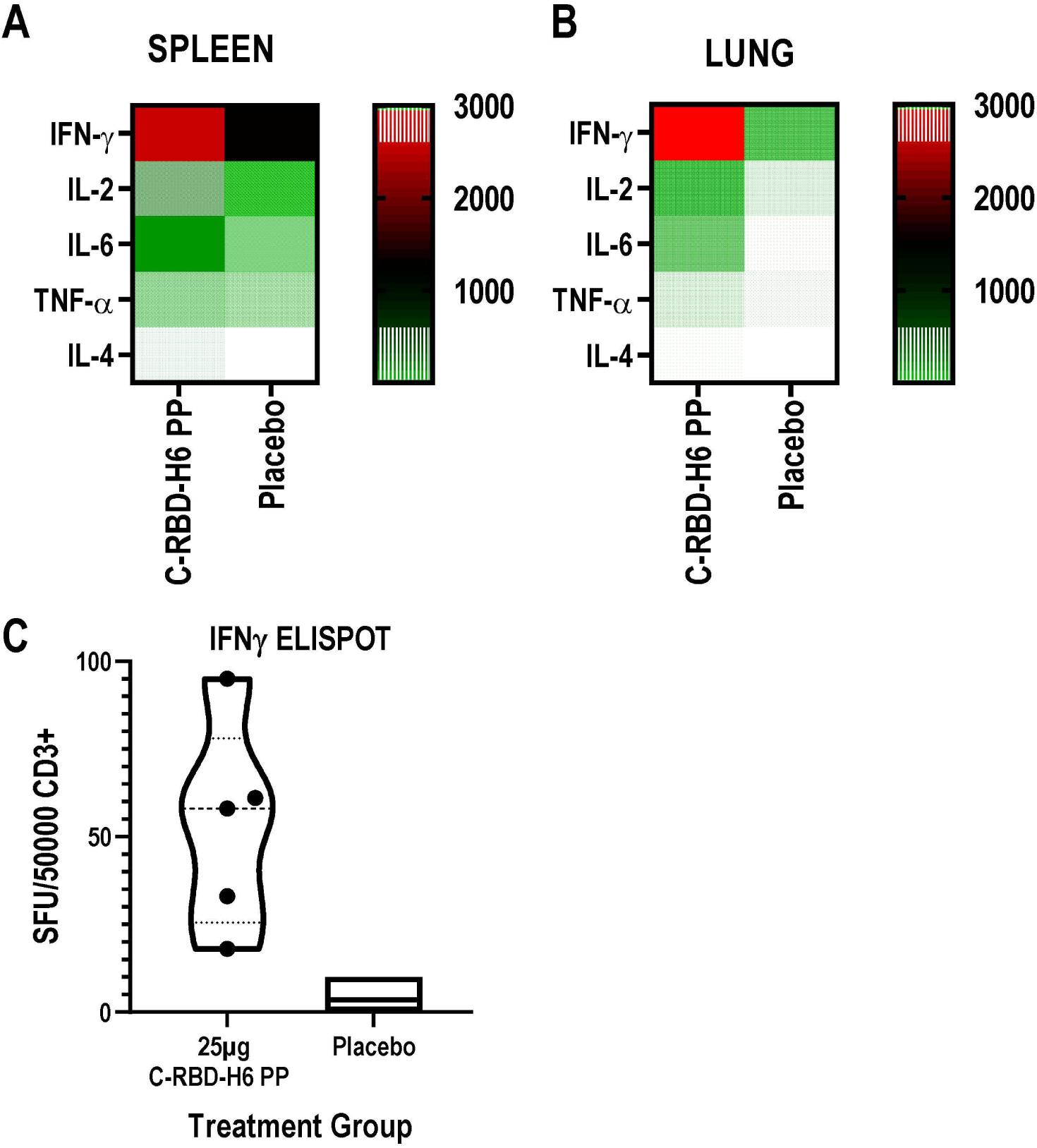
Heatmap of the cytokine response. Splenocytes (A) and Lung CD3+ enriched cells (B), after restimulation with C-RBD-H6 PP. Cells were obtained three months after the last immunization, from 4 to 5 mice per group that received three subcutaneous 25 µg doses. Non-stimulated controls were subtracted from re-stimulated samples.

## 4. Discussion

Despite the impressive development of prophylactic vaccines against COVID-19 and that several vaccines have reached the Emergency Use Authorization or Pharmaceutical Registry, there is not a sufficient supply of vaccines. In addition, the challenge of the continuous emergence of new mutant strains of the virus will require updating the antigen sequences included in the vaccines and administer booster doses to maintain the immunity of the population.

Results from this study demonstrate that it is possible to develop a vaccine candidate expressing RBD protein of SARS-CoV-2 using the yeast *P. pastoris*. The RBD was selected base on the state of the art evidence of the main contribution of the epitopes of RBD to the neutralization activity of the sera [13;26–28]

Moreover neutralizing antibody responses generated after immunization with Pfizer-BioNtech or Moderna vaccines of previously infected subjects, are mainly due to anti-RBD antibodies. The sera depletion of antibodies targeting the RBD abrogates sera neutralization capacity [29]

Optimal conditions for the production of a recombinant protein in *P. pastoris* expression system differ according to the target protein. Indeed, we were able to obtain 30-40 mg/L of the RBD with more than 98% of purity, close to the yield obtained in previous report by Arbeitman C.R., et al [8], an essential condition to develop a vaccine candidate. In addition to the protein yield, it is important the sugar composition. In *P. pastoris* the glycosylation pattern is characterized by the high mannose content. Mannosylation enhances activation of antigen-presenting cells like macrophages and dendritic cells, functioning as immunopotentiator while increases the antigen immunogenicity compared with its nonglycosylated counterparts [30]. The ionic interactions dependent on sugar composition also could help to stabilize RBD structure.

Our results on the affinity of the *P. pastoris* recombinant protein C-RBD-H6 PP by the receptor comply with those of other authors performing binding assays in BIACORE for the interaction pair RBD-ACE2 [31–33]. It means that probably the subunit vaccine based on C-RBD-H6 PP may elicit an effective antibody response against SARS-CoV-2, avoiding the virus entry and replication by blocking RBD-ACE2 interaction in real SARS-CoV-2 infection context.

The correct glycosylated protein during the transit in the endoplasmic reticulum and Golgi apparatus, go to the secretory pathway. If the protein is unable to fold properly is degraded in the cytosol. This mechanism guarantees that only properly folded proteins are secreted [8;34]. Our results demonstrated the presence of four intramolecular disulfide bonds identical to those present in the native RBD of SARS-CoV-2. These results as well as the secondary structure determined by CD spectroscopy confirm the right folding of the C-RBD-H6 PP.

Remarkably despite the known difference in the sugar composition of the protein expressed in *P. pastoris* compared to the expression systems in mammals, the antigenicity of the protein is similar both with polyclonal sera from mice and monkeys immunized with the recombinant protein, such as with human sera from individuals with natural coronavirus infection or vaccinated with the Pfizer-BioNTech or Sputnik V vaccines against SARS-CoV-2. The C-RBD-H6 PP protein also stimulated cellular response mediated by IFNγ secretion in lymphocytes isolated from convalescent subjects. The C-RBD-H6 PP protein also inhibited the RBD binding to the ACE2 receptor in a competitive ELISA regardless of whether the RBD receptor was obtained recombinant or it is found directly in the membrane of Vero E6 cells. In the same manner, the sera from mice, rats and NHP immunized with C-RBD-H6 PP protein inhibited the RBD-ACE2 receptor binding and neutralize the SARS-CoV-2 in microneutralization tests.

The need for the accelerated development of the vaccine candidate during the pandemic, led to the neutralizing activity of the NHP sera being evaluated just one week after the third dose, reaching titers comparable to the panel of convalescent sera used as a control. It is known that a long time for bleeding would lead to the maturation and selection of B cell clones producing high avidity antibodies and consequently higher neutralizing titers.

## Conclusions

Optimal conditions for the production of a recombinant protein in *P. pastoris* expression system differ according to the target protein. Indeed, we were able to obtain 30-40 mg/L of the RBD with more than 98% of purity.

Our ESI-MS results demonstrated the presence of four intramolecular disulfide bonds identical to those present in the native RBD of SARS-CoV-2, and the CD spectrum of the protein indicates the presence of well-packed aromatic and cysteine residues. These results together with the high receptor binding affinity in BIACORE assays confirm the right folding of the C-RBD-H6 PP protein.

The C-RBD-H6 PP antigenicity and the capacity of the sera from immunized mice, rats and NHP to inhibited the RBD-ACE2 receptor binding and neutralize the live virus infection to Vero E6 cells, points to the feasibility of the protein as a vaccine candidate.

## Data Availability

All relevant data are included in the manuscript or can be accessible upon request.

## Abbreviations

RBD: Receptor-binding domain
COVID-19: coronavirus disease 2019
SARS-CoV-2: severe acute respiratory syndrome coronavirus 2
ACE2: angiotensin-converting enzyme 2
PBMC: peripheral blood mononuclear cells
IL-2: Interleukin 2
IL-4: Interleukin 4
IL-6: Interleukin 6
TNFα: tumor necrosis factor-alpha
IFNγ: interferon gamma
NHP: non-human primates
CHO: Chinese hamster ovary cells
BHK21: baby hamster kidney cells
HEK293T: human embryonic kidney cells
IMAC: immobilized metal ion affinity chromatography
RP: reversed-phase chromatography
CD: circular dichroism
OD: optical density
VNT: viral neutralization titer

## Authors Contributions

GGN: provided original ideas and study concept and design, data curation, analysis and interpretation of data, and drafting, review and editing of the final version of the manuscript and studies supervision. MLF: contributed to analysis and interpretation of data, study design, review of the manuscript and studies supervision. LJGL: contributed to ESI-MS study design, acquisition of data, analysis and interpretation of data, and drafting and review of the manuscript. LAER, IAM and YRG: performed the ESI-MS studies, acquisition of data, analysis and interpretation of data. GCH: performed glycosylation studies. ACR: performed drafting and execution of Biacore studies. GMP: performed the protein purification studies. MPI and JZS: performed fermentation studies. GCS: provided original idea and drafting of the genetic construction, and performed structural analysis experiments by CD spectroscopy. AMMD: provided original ideas and performed the genetic construction and review of the manuscript. and DGR: performed the genetic construction and protein expression experiments. MBR: provided original ideas, study designs, analysis and interpretation of data, drafting and review of the manuscript, graphic and statistical processing and execution of antigenicity, immunogenicity and cellular response studies. IGM and CCHA: performed antigenicity, immunogenicity studies and analysis of cellular response in mouse, rat, and NHP. OCS: study design and supervision of the microneutralization experiments. GLP: contributed to analytical procedures and donor patients selection and evaluation of the immunological and functional response. JVH, EMD, EPV and MAA: study concept and supervision.

## Disclosure

MLF, MBR, AMMD, DGR, ACR, GCS, GMP, EPV, MAA and GGN are co-authors of the Center for Genetic Engineering and Biotechnology patent application comprising the C-RBD-H6 PP protein as a vaccine antigen against SASR-CoV-2.

All authors approved integrally the final article.

## Acknowledgment

The authors acknowledge Dr. Gabriel Padron and Dr. Diana Garcia del Barco Herrera for their writing assistance. We also thank Dr. Gertrudis Rojas, Dr. Tays Hernández and Dr. Belinda Sanchez from the Center for Molecular Immunology for the recombinant mFc-RBD and hFc-RBD chimeric proteins.

## Appendix. Other co-authors in the Study

### Center for Genetic Engineering and Biotechnology

Isela María Garcia Tamayo

Yahima Chacón Quintero

Ricardo Ulises Martinez Rosales

Dionne Casillas Casanova

Ailyn de la Caridad Ramón Sánchez

Lizet Aldana Velazco

Jorge Castro Velazco

### Civilian Defense Scientific Research Center

Juliet María Enriquez Puertas, M.Sc.

Mireida Rodríguez Acosta, M.D., Ph.D.

Enrique Noa Romero, Ph.D.

Nibaldo Luis González Sosa, M.Sc.

Marta Dubed Echevarría, M.Sc.

