## Supplemental File S1 for "The SARS-CoV-2 receptor-binding domain expressed in Pichia pastoris as a candidate vaccine antigen"

**Supplement S1**

Protein sequence

NWSFFSNIGGSSGGSNITNLCPFGEVFNATRFASVYAWNRKRISNCVADYSVLYNSAS
FSTFKCYGVSPTKLNDLCFTNVYADSFVIRGDEVRQIAPGQTGKIADYNYKLPDDFTGC
VIAWNSNNLDSKVGGNYNYLYRLFRKSNLKPFERDISTEIYQAGSTPCNGVEGFNCYF
PLQSYGFQPTNGVGYQPYRVVVLSFELLHAPATVCGPKKGGSGGSSSSSSSSSSIEH
HHHHH
